# ACE inhibition increases Alzheimer’s disease risk by promoting tau phosphorylation

**DOI:** 10.1101/2025.08.11.25333412

**Authors:** Min Seo Kim, Sunyoung Park, Jong Ho Kim, Woojae Myung, Minku Song, Ron Do, Kwangsik Nho, Eunae Kim, Seonghoon Hwang, Zhi Yu, Pradeep Natarajan, Hee Jin Kim, Hyo Jin Son, Woong-Yang Park, Yang Sui, Akl C Fahed, Patrick T. Ellinor, Muhammad Ali, Katherine Gong, Carlos Cruchaga, Dong Keon Yon, Jinsoo Seo, Ju-Young Shin, Dong-Gyu Jo, Hong-Hee Won

## Abstract

Reportedly, over 60% of individuals in the USA aged 65 or older take antihypertensive medications, making it crucial to evaluate their potential impact on dementia. Alzheimer’s disease (AD), the most prevalent form of dementia, develops insidiously over decades, effectively precluding clinical trials of antihypertensive drug effects on AD risk. Through a triangulation approach integrating large-scale human genetics, population-based study, and rigorous experimental models, we identified that angiotensin-converting enzyme (ACE) inhibitors were associated with increased AD risk, with no significant associations observed for other antihypertensive classes, including angiotensin II receptor blockers and calcium channel blockers. Multi-omics Mendelian randomization analyses revealed that genetically proxied reductions in *ACE* expression and ACE protein levels in serum, cerebrospinal fluid, and brain tissues were causally associated with increased AD risk via exacerbated tau phosphorylation. A cohort study of 338,645 UK Biobank participants, followed for more than 10 years, also found an association between genetically predicted low serum ACE levels and the increased incidence of AD. Experimental validation using P301S tau transgenic mice and human iPSC-derived neurons confirmed that pharmacological ACE inhibition intensified tau phosphorylation and aggregation, cognitive impairment, neuroinflammation, and glial activation, predominantly in the hippocampal region. Multiple lines of evidence established a specific link between ACE inhibition and tau-driven neurodegeneration, underscoring the importance of carefully tailored antihypertensive strategies to prevent dementia risk, and identifying ACE as a viable therapeutic target for AD and other tau-related neurodegenerative conditions.

## Introduction

Given that Alzheimer’s disease (AD) is particularly prevalent in older people with hypertensive comorbidities, the potential impact of antihypertensive drugs on dementia represents an important clinical concern. In the USA, more than 60% of individuals aged over 65 years are on antihypertensive medications (*1*). However, the association between antihypertensive agents and dementia risk remains controversial.

This association is challenging to determine using conventional clinical methodologies, owing to long asymptomatic progression and late manifestation of AD. Although randomized clinical trials (RCTs) represent the gold standard for establishing causality, the long latency period inherent in AD necessitates impractically extended follow-up durations of 10–20 years. The mean follow-up duration of RCT investigating this association is 4.1 years (*2*), which is significantly shorter than the timeframe required to reliably assess chronic neurodegenerative conditions. Conversely, prospective cohort studies, despite their capability to track long-term outcomes, are intrinsically vulnerable to confounding factors and can ascertain associations rather than causation (*3, 4*). To resolve these methodological constraints and elucidate the causal association between antihypertensive agents use and AD risk, we designed a triangulation approach that synthesized findings from large-scale human genetics, population level analyses, and mechanistic studies using both *in vitro* assays and *in vivo* models.

We first conducted a Mendelian randomization (MR) analysis, which used genetic variants that were randomly allocated during meiosis as instrumental variables. This approach conceptually emulates the random allocation of participants in an RCT (*3, 5*), offering a natural experiment for causal inference and minimizing confounder risk (*6, 7*). Thus, an MR study provides a unique opportunity to complement clinical studies (*8–10*), and is increasingly used to evaluate the efficacy and safety of drugs (*11*).

In a hypothesis-free, large-scale human genetics study, we screened 23 drug target genes from 10 antihypertensive drug classes, including angiotensin-converting enzyme (ACE) inhibitors, angiotensin II receptor blockers (ARBs), calcium channel blockers (CCBs), and diuretics, for their association with AD risk. The initial screening indicated a strong association between ACE inhibitors and AD. Subsequently, we performed comprehensive mechanistic studies using both *in vitro* and *in vivo* models to validate and elucidate the biological mechanisms underlying the observed association between ACE inhibition and AD pathology.

## Results

### Large-scale human genetics study identifies that ACE inhibition potentially increases the risk for AD

We first conceptualized the strengths of MR compared with those of RCTs (Fig. 1), emphasizing the capability of MR to address the methodological challenges inherent to conventional clinical research, particularly pertinent to our research question. We began our investigation with a multi-omics MR study, employing genetic instruments (GIs) associated with drug target gene expression (expression quantitative trait loci; eQTLs) and protein abundance (protein quantitative trait loci; pQTLs) (Fig. 1, Fig. S1). The rigorous GI selection process shown in Fig. S1 was used to identify 23 drug target genes from 10 antihypertensive drug classes. Among them, only genetic variants associated with decreased *ACE* expression was significantly associated with AD at the Bonferroni threshold of *P<*8.92 × 10^−4^ (Fig. 2B). A 1-standard deviation (SD) decrease in blood *ACE* expression (mimicking ACE inhibitor) was associated with 3.71 mmHg decrease in systolic blood pressure (SBP) as expected, and a higher risk of AD (odds ratio (OR), 2.00; 95% confidence interval (CI), 1.47–2.70; *P*, 7.51 × 10^−6^) (Fig. 2B). Details of the summary data-based MR (SMR) results for the analysis of the expression of 23 drug target genes and AD are provided in Tables S6–7. In addition to serum *ACE* expression, decreased *ACE* expression levels in the cerebellum, frontal cortex, and nerve tissues were significantly associated with an increased AD risk (Fig. 2C).

**Fig. 1.**
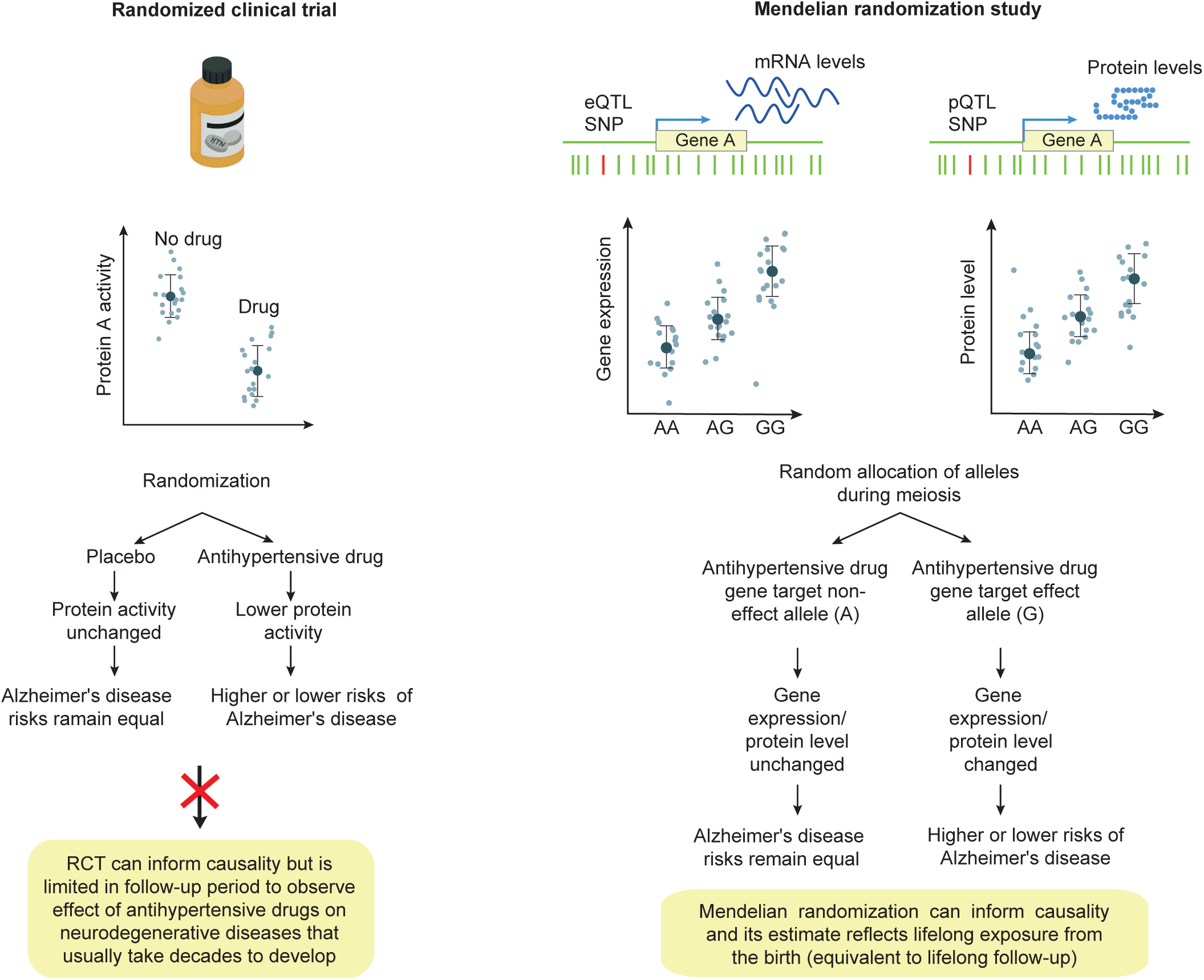
Comparative schematic of a randomized controlled trial (RCT) and Mendelian randomization (MR) approaches for pharmacovigilance. Single-nucleotide polymorphisms (SNPs) associated with antihypertensive drug target gene expression (expression quantitative trait loci [eQTL]) and target protein levels (protein quantitative trait loci [pQTL]) were used as instrumental variables for drug exposure. MR leverages the random allocation of risk alleles at conception to infer causality and effectively emulates the random allocation of participants in an RCT.

**Fig. 2.**
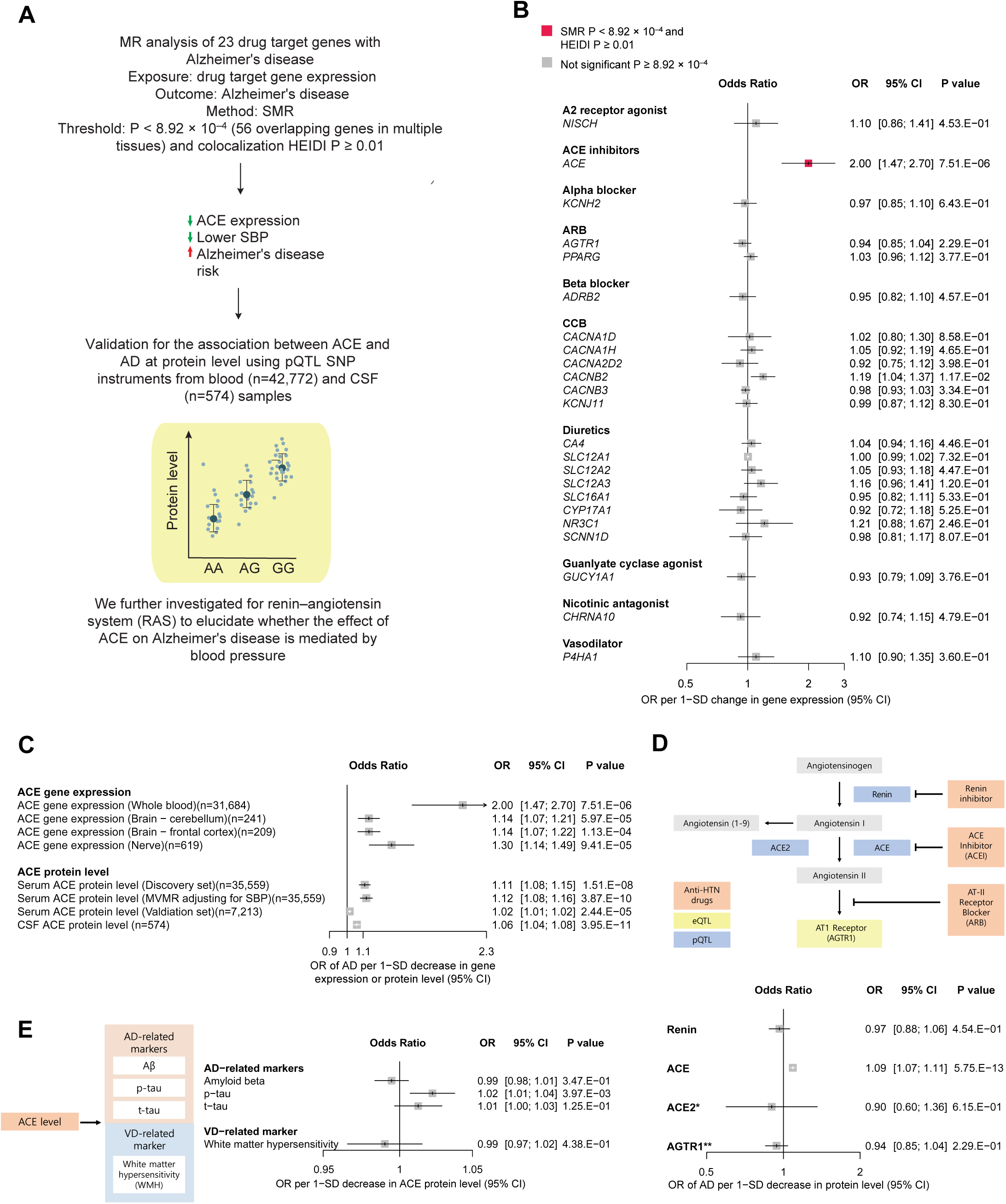
Genetic association between antihypertensive drug targets and AD in large-scale human genetics analyses. (**A**) Schematic of the human genetic study and validation. (**B**) Causal association between drug target gene expression and AD. Since each antihypertensive drug acts as either agonist or antagonist on drug targets, the odds ratio (OR) was harmonized to reflect the drug’s actual effect on target genes. Therefore, OR greater than 1.00 indicates an increased risk of disease by the use of antihypertensive drugs. The drug effect on the target gene is identified from DrugBank and ChEMBL. Except for *AGTR1* (nerve tissue), all gene targets in the forest plot were derived from whole blood. (**C**) Causal association of *ACE* expression and protein levels with AD in multiple tissues. To reflect the antagonist effect of the ACE inhibitor on ACE levels, the OR per 1-standard deviation (SD) decrease in ACE gene expression or protein levels was assessed. (**D**) Causal associations of up/downstream enzymes of ACE in the renin-angiotensin system with AD were analyzed to evaluate the blood pressure-mediating effect. *No *cis*-pQTL SNP was available for ACE2; therefore, ACE2 *trans*-pQTL SNP was used instead. **Since there was no available pQTL for AGTR1 (drug target of ARB), expression QTL was used instead. (**E**) Causal associations of serum ACE levels with AD- and vascular dementia-related markers were analyzed. HEIDI, Heterogeneity in dependent instruments; pQTL, protein quantitative trait loci; SNP, single-nucleotide polymorphism; SMR, summary data-based Mendelian randomization; SBP, systolic blood pressure; CSF, cerebrospinal fluid; AD, Alzheimer’s disease. OR, odds ratio; CI, confidence interval; SD, standard deviation; ACE inhibitor, angiotensin-converting enzyme inhibitor; ARB, angiotensin II receptor blocker; CCB, calcium channel blocker; p-tau, phosphorylated tau; t-tau, total tau.

To further confirm the genetic association between ACE and AD, we validated the association between ACE and AD at the protein level using pQTL-based single nucleotide polymorphism (SNP) derived from blood (n=42,772) and cerebrospinal fluid (CSF; n=574). Employing multiple MR analytic methods other than SMR, including inverse variance-weighted, MR Egger, weighted median, and MR-PRESSO, we consistently observed that genetically predicted lower ACE protein levels in the serum were significantly associated with increased AD risk in both the discovery and validation cohorts, even after adjusting for SBP (Fig. 2C, Fig. S1; Tables S12-13). Similarly, decreased ACE protein levels in the CSF were associated with elevated AD risk (OR, 1.06; 95% CI, 1.04–1.08; *P* = 3.95 × 10^−11^; Fig. 2C, Table S11).

Importantly, this association appeared to be ACE-specific, as genetic variations affecting other enzymes in the renin-angiotensin system (RAS) were not significantly associated with AD risk (Fig. 2D, Tables S10, S14-S15). This specificity suggests an underlying mechanism independent of BP regulation. To explore the potential non-BP dependent mechanisms, we performed MR analysis linking ACE to AD and vascular dementia (VD) biomarkers. Lower serum ACE levels were significantly associated with increased CSF phosphorylated tau (p-tau) levels (OR, 1.02; 95% CI, 1.01–1.04; P = 3.97 × 10^−3^; Fig. 2E, Table S17), implicating tau phosphorylation as a potential BP-independent pathogenic pathway linking ACE inhibition to AD. A full description of the findings from human genetics study is provided in the supplementary text.

### ACE inhibition is associated with increased risk of AD in 338,645 UK Biobank participants

To further evaluate the clinical implications of ACE inhibition on AD risk at the population level, we conducted a cohort study in 338,645 UK Biobank participants. Genetically predicted serum ACE level was calculated using polygenic risk score (PRS) for serum ACE levels. The low ACE PRS group includes those with genetically predicted low serum ACE levels, emulating ACE inhibition. Compared with the high PRS group (those with genetically high serum ACE levels), the OR of AD was 1.21 (95% CI, 1.06–1.38; P, 6.12 × 10−3) for the intermediate PRS group and 1.26 (95% CI, 1.07–1.48; P, 5.06 × 10−3) for the low PRS group in the overall population (Fig. 3). This association was slightly stronger among APOE4 allele carriers than non-carriers (Fig. 3).

**Fig. 3.**
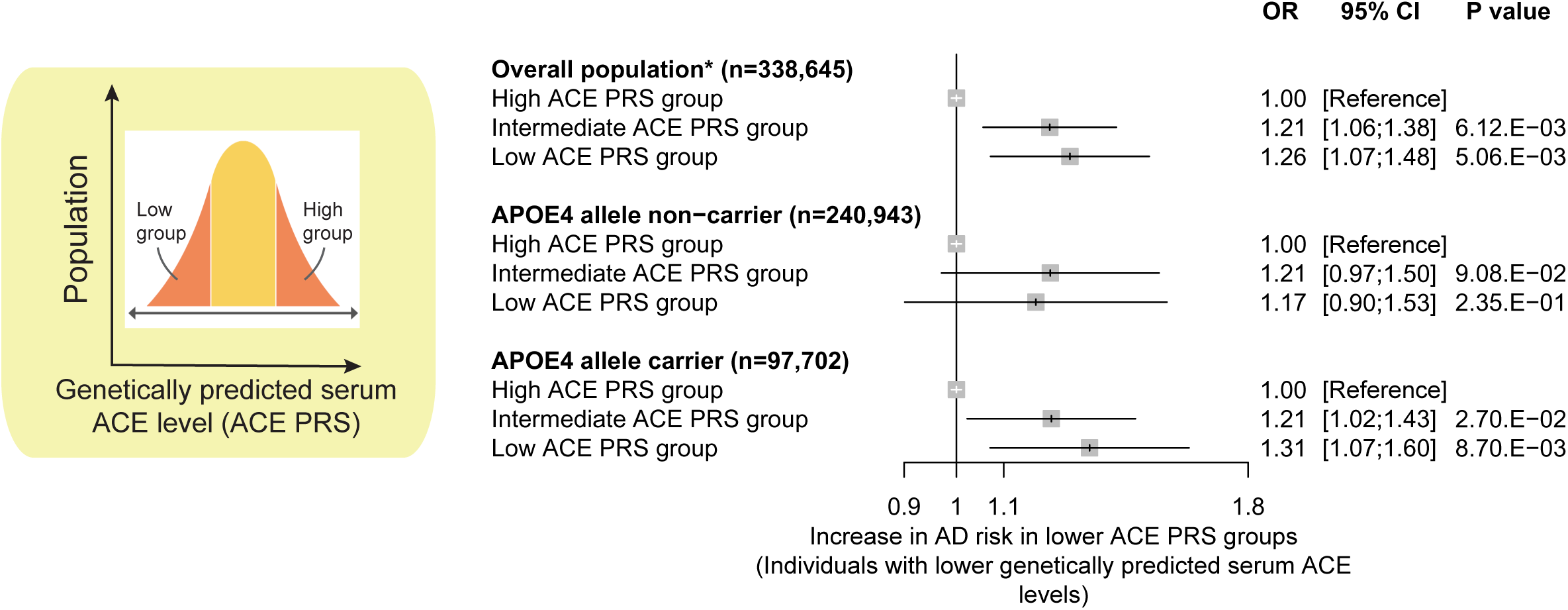
Genetically predicted low serum ACE level is associated with increased risk of Alzheimer’s disease (AD) in 338,645 UK Biobank participants. Association of polygenic risk score (PRS) for serum ACE levels and AD in unrelated UK Biobank participants (n=338,645). The low ACE PRS group includes those with genetically predicted low serum ACE levels. The top 20% was categorized into the high ACE PRS group, 60% into the intermediate ACE PRS group, and the bottom 20% into the low ACE PRS group. Lower PRS infers a lower genetically predicted ACE levels and thus odds ratio (OR) greater than 1.00 emulates the effect of ACE inhibition. *The model was adjusted for age, sex, education, smoking, alcohol consumption, depression, diabetes mellitus, hypertension, body mass index, APOE ε4 (APOE4) risk allele count (0, 1, 2), genotype array, and the first 10 principal components of ancestry.

### Pharmacologic ACE inhibition exacerbates tau pathology and memory deficits in P301S tau transgenic mice

To experimentally validate the role of ACE inhibition in tau pathology, as suggested by our genetic findings, we conducted a comprehensive series of *in vitro* and *in vivo* experiments. Initially, we assessed tau aggregation in Tau-BiFC cells treated with ACE inhibitors (ramipril and enalapril) or ARBs (telmisartan, losartan, olmesartan). Treatment of ACE inhibitors significantly increased tau aggregation, as indicated by the enhanced fluorescence intensity, whereas ARBs did not induce this effect (Fig. S4, A and B), supporting a specific role for ACE inhibition in promoting tau aggregation.

Subsequently, we investigated the effects of ACE inhibition on cognitive function in P301S tau transgenic mice. Four-month-old mice received daily oral administration of a BBB-penetrating ACE inhibitor (ramipril), a non-penetrating ACE inhibitor (enalapril), or an ARB (losartan) for four months (Fig. 4A and Fig. S5A). After two months, spatial learning and memory were evaluated using the Morris water maze (MWM). While vehicle-treated P301S (P301S Veh) mice exhibited no significant spatial memory impairments, ramipril-treated P301S (P301S Rami) mice demonstrated pronounced cognitive deficits compared to wild-type (WT) controls (Fig. 4B). To assess the sustained effects of ACE inhibition on cognitive function, treatment continued for two additional months, followed by a second MWM assessment (Fig. 4A, Fig. S5B). The ramipril-treated mice consistently exhibited spatial memory deficits (Fig. 4C, Fig. S5C). Cognitive impairment specific to ACE inhibitors was confirmed using the novel object recognition (NOR) test (Fig. S5D–F).

**Fig. 4.**
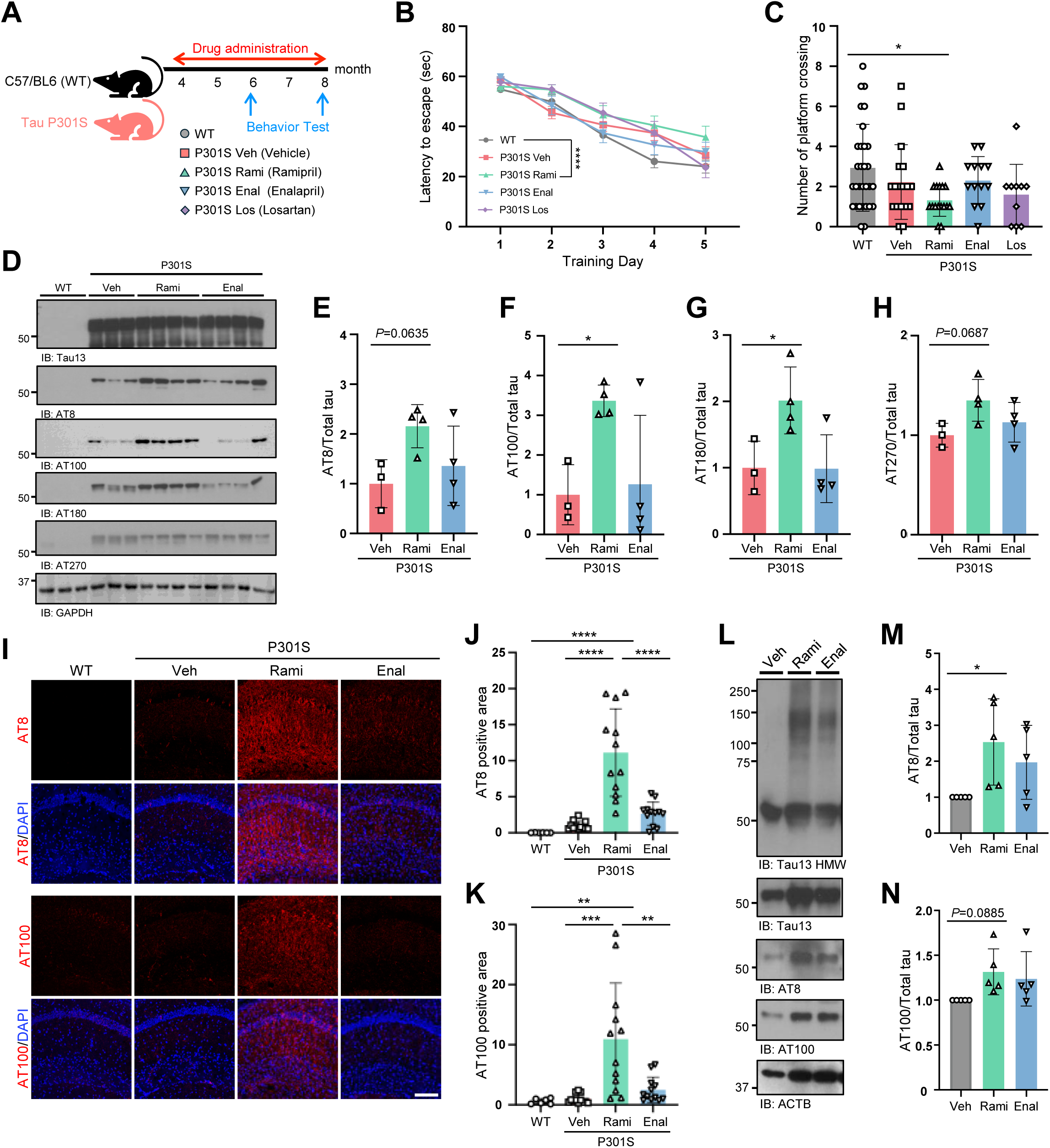
ACE inhibitors exacerbate cognitive impairment and tau pathology. (**A**) Schematic of the experimental design. ACE inhibitors (ramipril, enalapril) and an angiotensin receptor blocker (ARB; losartan) were administrated orally. Group sizes: WT (*n* = 30), P301S Vehicle (*n* = 18), P301S Ramipril (Rami; *n* = 16), P301S Enalapril (Enal; *n* = 13), P301S Losartan (Los; *n* = 10). (**B**) Escape latency during training in the Morris water maze (MWM) at 6 months of age. (**C**) Number of entries into platform zone in the MWM test at 8 months of age. (**D**) Immunoblot analysis of total tau and phosphorylated tau (p-Tau) in hippocampal lysates. (**F-K**) Quantification of p-Tau species: (**E**) AT8, (**F**) AT100, (**G**) AT180, (**H**) AT270. (**I**) Immunohistochemical analysis of brain sections stained with AT8 and AT100. (**J** and **K**) Quantification of (**J**) AT8 and (**K**) AT100 positive area. (**L**) Immunoblot analysis of total tau and p-Tau in human iPSC-derived cortical neurons. (**M** and **N**) Quantification of p-Tau in human iPSC-derived cortical neurons. Statistical analyses: **B** two-way ANOVA with mean ± SEM (*P < 0.05, ****P < 0.0001); **C, J**, and **K**, one-way ANOVA with Tukey’s multiple comparisons test, mean ± SD (*P < 0.05, **P < 0.01, ***P < 0.001, ****P < 0.0001); **E-F, M**, and **N**, one-way ANOVA with Dunnett’s multiple comparisons test, mean ± SD (*P < 0.05). Scale bar, 40µm.

To determine whether these cognitive impairments correlated with increased tau pathology, we analyzed tau phosphorylation in cortical and hippocampal tissues. Ramipril-treated P301S mice exhibited significantly higher levels of tau phosphorylation than vehicle- or enalapril-treated mice, suggesting a specific effect of BBB-penetrating ACE inhibitors on tau phosphorylation (Fig. 4D–H, Fig. S6). Immunohistochemical analysis further revealed markedly elevated phosphorylated-tau immunoreactivity in the hippocampi of ramipril-treated mice than other groups (Fig. 4I–K, Fig. S7).

To extend these observations to human neuronal systems, we generated cortical neurons from induced pluripotent stem cells (iPSCs) derived from a patient with *APOE4/4* genotype (Fig. S8A). Treatment with ACE inhibitors, but not ARBs, significantly increased tau phosphorylation and increased high-molecular-weight tau levels, indicating increased tau aggregation (Fig. 4L–N and Fig. S8B–C). Notably, tau aggregation increased in neurons derived from *APOE4/4* iPSCs and *APOE3/3* iPSCs (Fig. S8B and C). Furthermore, cortical neurons differentiated from human embryonic stem cells exhibited elevated tau phosphorylation and aggregation following treatment with ACE inhibitors (Fig. S8D–F). Collectively, these findings demonstrate that pharmacological ACE inhibition exacerbates tau phosphorylation and aggregation, thereby contributing to the cognitive deficits observed in both human neuronal models and P301S tau transgenic mice.

### ACE inhibition promotes inflammatory gene expression and glial activation in P301S tau mice

Previous studies have shown that pathological tau induces neuroinflammation (*12–14*). To investigate whether ACE inhibition exacerbates this neuroinflammatory response, we conducted bulk RNA sequencing of the cortical tissue from P301S tau mice treated with ramipril or vehicle. Ramipril treatment significantly altered the expression of 412 genes compare to vehicle-treated controls (Fig. 5A and Table S20). Gene Ontology (GO) enrichment analysis revealed that the upregulated genes were primarily associated with inflammatory signaling pathways (Fig. 5B). In particular, immune response-related pathways, including type II interferon signaling and cytokine-mediated T cell activation, were correlated with upregulated genes, which is consistent with previous studies (*15, 16*). Conversely, the downregulated genes were enriched in neuronal and astrocytic developmental processes (Fig. 5C), indicating disrupted neuroglial homeostasis upon ACE inhibition. These transcriptomic data imply that ACE inhibition accelerates cognitive decline by heightening inflammatory responses and impairing neuroglial function.

**Fig. 5.**
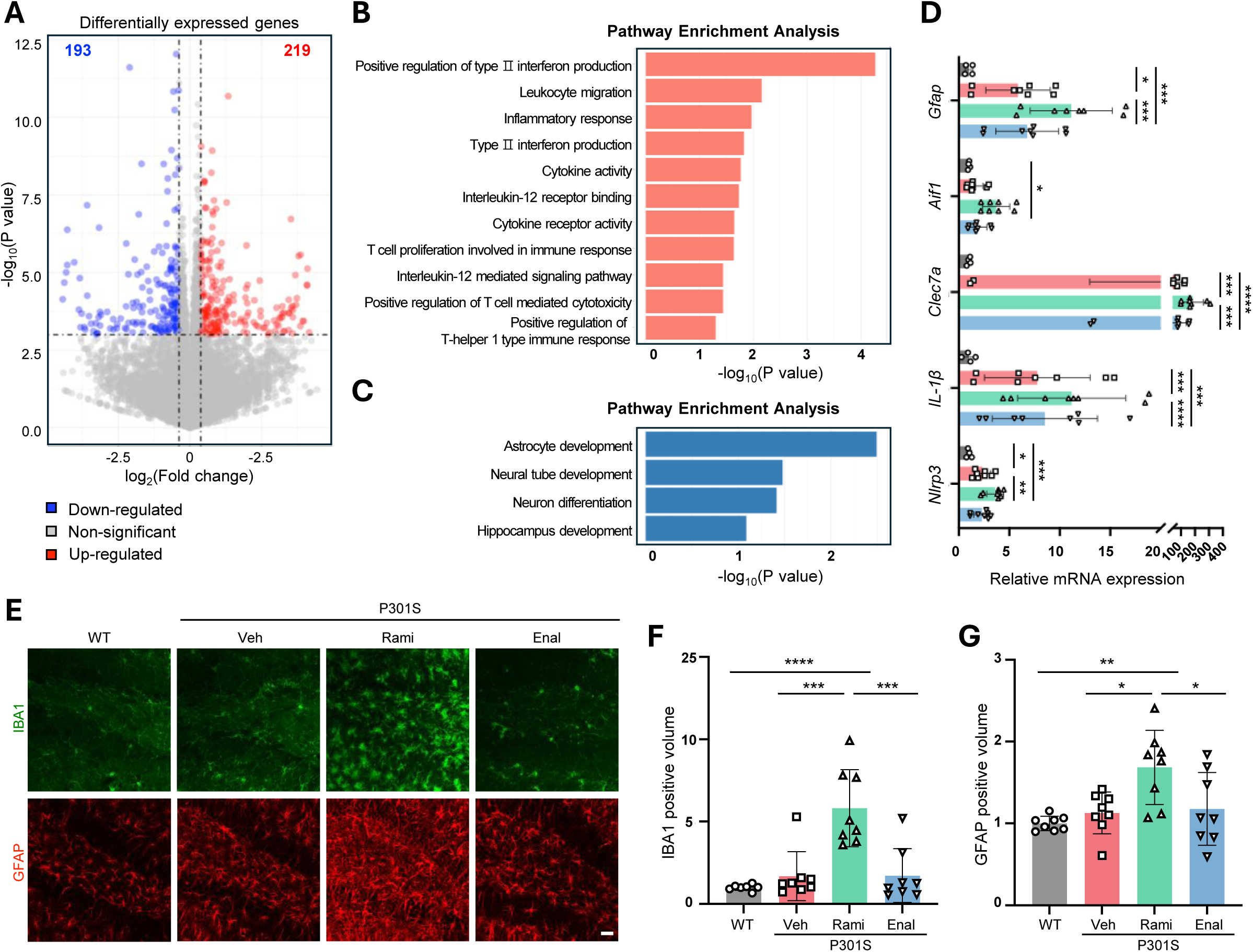
ACE inhibitors induce inflammatory responses in P301S mice. (**A**) Volcano plot showing differentially expressed genes (DEGs) in cortical tissue of P301S mice treated with ramipril (P301S Rami) versus vehicle (Veh). (**B** and **C**) Gene Ontology (GO) classification of DEGs in P301S Rami mice: (**B**) upregulated pathways and (**C**) downregulated pathways. (**D-H**) mRNA expression levels of inflammatory markers in hippocampal lysates from mice treated with ACE inhibitors: (**D**) *Nlrp3*, *Il1b*, *Clec7a*, *Aif1* (Iba1), *Gfap*. (**E**) Immunohistochemical analysis of hippocampal sections stained with Iba1 and GFAP. (**F** and **G**) Quantification of (**F**) Iba1-positive volume. (**G**) GFAP-positive volume. Scale bar, 5µm Data are shown as mean ± SD. Statistical significance was determined by one-way ANOVA followed by Tukey’s multiple comparisons test (*P < 0.05, **P < 0.01, ***P < 0.001, ****P < 0.0001). P301S vehicle (Veh), P301S ramipril (Rami), P301S enalapril (Enal). Scale bar, 20µm.

Owing to our transcriptomic evidence of inflammation, we next evaluated glial activation at the tissue level. Ramipril-treated P301S mice displayed significantly elevated expression of genes related to innate immune response (*Nlrp3*, *Il-1β*, and *Clec7a*), microglial activation (*Clec7a* and *Aif1*), and astrocyte reactivity (*Gfap*) in both cortical and hippocampal tissues (Fig. 5D, Fig. S9). Immunohistochemical analysis further revealed substantial increases in microglial (Iba1-positive) and astrocytic (GFAP-positive) activation, particularly in the hippocampal regions of ramipril-treated mice than in vehicle-treated controls (Fig. 5E–G, Fig. S10A). Although most pronounced in the hippocampus, significant microglial activation was also observed in cortical regions (Fig. S10B–D), indicating widespread neuroinflammatory responses induced by ACE inhibitor-associated tauopathy.

To determine whether the observed glial activation was directly induced by ACE inhibitor or secondary to exacerbated tau pathology, mouse primary microglial cultures were treated with ramipril or enalapril. These ACE inhibitors significantly altered the expression of inflammatory genes *in vitro* (Fig. S11), suggesting that the glial activation observed in vivo may predominantly result from enhanced tau pathology rather than a direct pharmacological effect. Collectively, these findings demonstrate that ACE inhibition potentiates tauopathy-associated neuroinflammation by amplifying immune transcriptional responses and subsequent glial activation.

## Discussion

Through an integrative approach combining large-scale human genetics, population-based analyses, and mechanistic validation *in vitro* and *in vivo*, our study revealed that ACE inhibition increased risk of AD through exacerbating tau phosphorylation and aggregation. Our human genetic analyses consistently demonstrated that reduced *ACE* expression and protein levels in serum, CSF, and brain tissues were causally associated with an elevated AD risk. These findings were substantiated by behavioral and histopathological assessments in P301S tau transgenic mice and human iPSC-derived cortical neurons, where ACE inhibition enhanced tau aggregation and subsequent tauopathy-associated neuroinflammatory responses. Furthermore, a longitudinal epidemiological analysis of UK Biobank data demonstrated an elevated AD risk associated genetically predicted low serum ACE level, providing population-level validation of our human genomic and preclinical results.

The involvement of ACE in AD pathophysiology has garnered increasing attention, predominantly focused on amyloid-β (Aβ) clearance mechanisms. Previous research demonstrated that microglial ACE enhances Aβ phagocytosis via ACE catalytic activity and SYK signaling pathways (*17*), with further studies reporting ACE-mediated enhancement of Aβ clearance (*18–20*). Macrophage-specific ACE overexpression also reduced pathological Aβ accumulation and preserved synaptic function in mouse models (*21*). Despite some evidence implicating ACE in Aβ metabolism, its relationship with tauopathy remains notably understudied, representing a critical gap in our current understanding of the comprehensive role of ACE in AD pathology. To date, only one study reported reduced tau phosphorylation and total tau levels following lisinopril treatment in mice, suggesting a potential therapeutic benefit of ACE inhibitors in AD (*22*). In contrast, our comprehensive multi-model approach demonstrated the opposite, in which increased tau phosphorylation and aggregation were observed following ACE inhibition, implying an adverse effect of ACE inhibitors on AD. These data add important mechanistic insights into the role of ACE beyond Aβ metabolism, highlighting tau phosphorylation as a novel and significant pathway implicated in ACE-mediated AD risk. Altogether, ACE inhibition likely contributes to the pathologies associated with both Aβ and tau. This dual function highlights its pivotal role in the pathophysiology of AD, positioning ACE as a strong candidate for therapeutic intervention.

The association between prescribing antihypertensive agents and dementia risk holds significant clinical relevance given the high prevalence of these drugs use in aging populations. Our findings indicating detrimental effects of ACE inhibitors on AD risk initially appear inconsistent with prior meta-analysis of RCTs suggesting protective roles of antihypertensive drugs in dementia (*2*). This discrepancy can be attributed to several factors. First, Hughes et al. conducted a meta-analysis of RCTs using all-cause dementia, a composite of VD and AD, as the outcome, making it difficult to isolate the specific effects of antihypertensive drugs on AD alone (*2*). Indeed, our subgroup reanalysis of the RCTs that focused solely on AD outcomes produced neutral results (Fig. S3), consistent with our MR findings, which showed no association between most antihypertensive drug classes–except ACE inhibitors–and AD (Fig. 2B). Thus, our findings are not necessarily discordant with previous clinical evidence; rather, they offer complementary insights. Secondly, the cerebrovascular benefits of BP reduction via antihypertensive therapy may mask or offset the detrimental effects of ACE inhibitors, resulting in neutral or protective outcomes in clinical studies. If this mechanism is validated, use of non-ACE-targeting antihypertensive agents should be preferred to provide optimal cerebrovascular protection without exacerbating tau-related pathology, potentially representing a superior clinical strategy for reducing AD risk.

It may be premature to categorically avoid ACE inhibitors; however, given their status as one of the world’s most prescribed drug classes (*23*), a cautious approach is justified. For individuals with genetic or clinical susceptibility to AD, or younger patients requiring long-term antihypertensive treatment, ARBs or other antihypertensives may be prudent options with a favorable benefit–risk balance. If ACE inhibitors are clinically indicated, preference might be given to non-BBB-penetrant formulations as enhanced tau phosphorylation with BBB-penetrating ACE inhibitors over non-penetrating formulations was observed in our mouse model. Our finding aligns with a large cohort study by Newby et al. of over 350,000 individuals, which found centrally active ACE inhibitors were associated with higher dementia risk than their non-centrally active counterparts (*24*). Additionally, based on mechanistic inference, careful use of ACE inhibitors may be advisable for other tau-related neurodegenerative disorders, such as frontotemporal dementia, progressive supranuclear palsy, Pick’s disease, and corticobasal degeneration (*25*), which warrants further investigation.

Our study has several limitations. First, the absolute effect sizes derived from the MR analyses (such as effect sizes at the gene expression and protein levels in Fig. 2C) should be interpreted with caution. MR is primarily intended to test causality rather than precisely quantify the effects. Therefore, comparisons of effect magnitudes across different outcomes are not meaningful (*5, 26*). Secondly, MR is based on genetic variants inherited at conception and assumes lifelong exposure rather than an acute effect (*27*). Therefore, the potential adverse effects of ACE inhibitors should not be broadly extrapolated to short-term or intermittent use. As our analyses could not determine the precise exposure durations needed to induce adverse effects, future studies should investigate pharmacological parameters, such as dose–response relationships, drug metabolism, and interactions with concomitant medications, to clarify the exposure thresholds and kinetics underlying the observed risks. Thirdly, the P301S tau transgenic mouse model employed in our study recapitulates tauopathy through mutant tau overexpression but does not include Aβ pathology, a hallmark of human AD. Thus, although valuable for mechanistic insights into tau pathology, this model incompletely captures the multifactorial complexity of human AD, potentially limiting its translational applicability. To address this limitation, we complemented our animal model findings with human-relevant iPSC-derived neurons that demonstrated increased tau phosphorylation and aggregation after ACE inhibitor treatment. Additionally, our epidemiological analyses provided real-world validation by demonstrating an increased risk of AD associated with genetically proxied ACE inhibition, thus enhancing the translational potential of our findings.

Our findings indicate that ACE inhibition may potentiate tau pathology and cognitive impairment, whereas other antihypertensive classes, including ARBs and CCB, do not appear to increase AD risk. This multimodal triangulation of evidence across large-scale human genetics, cohort study, and experimental models highlights a robust framework for pharmacovigilance and provides critical insights into complex drug–disease interactions that are not easily captured by clinical trials.

## Methods

### Human Genetics Study

#### Study Design and Data Sources

All studies providing data for this study had relevant institutional review board approval as per the Declaration of Helsinki, and informed consent was obtained from all participants. The UK Biobank study was approved by the National Information Governance Board for Health and Social Care and the North West Multicenter Research Ethical Committee (11/NW/0382). All participants provided informed consent. This study was conducted using the UK Biobank Resource (Application No. 33002). We followed the Strengthening the Reporting of Observational Studies in Epidemiology - Mendelian Randomization (STROBE-MR) reporting guidelines (*28*). Summarized genetic association data of AD were obtained from previous genome-wide association study (GWAS) meta-analyses (Table S1) (*29–31*), and details of the samples and genome-wide association analytic workflow have been provided previously (*29–31*). The overall study design and analytical workflow are shown in Fig. 1 and Fig. S1, respectively.

We obtained data on common (minor allele frequency [MAF] > 1%) single-nucleotide polymorphisms (SNPs) associated with gene expression in the blood from the eQTLGen consortium (n = 31,684, Table S2) (*32*), and included only *cis*-expression quantitative trait loci (*cis*-eQTL) (distance between SNPs and a gene within 1 Mb). The *cis*-eQTL for 12 different brain regions and two nerve tissues were retrieved from the latest version of the Genotype-Tissue Expression (GTEx) database (v8), with sample sizes ranging from 176 to 619 (Table S2) (*33*). We also used protein quantitative trait loci (pQTL) for analysis, since consolidating evidence from eQTL and pQTL has a synergistic resolution for drug target investigations (*34*). Genetic data for blood protein levels from 35,559 Icelanders and 7,213 European participants of the Atherosclerosis Risk in Communities (ARIC) study were obtained, and *cis*-pQTLs were used for analysis (Table S3) (*35, 36*). The *cis-*pQTL was limited to SNPs within 1 Mb on either side of the protein-encoding gene for Icelander pQTL data in accordance with *cis-*eQTL data from eQTLGen and GTEx; yet, a ±500 Kb window size was exceptionally used for ARIC pQTL as per the discrete definition of the study (*36*). The eQTL and pQTL data are on the scale of one 1-standard deviation (SD) change in gene expression/natural log-transformed protein levels for each additional effect allele.

#### Drug Target Gene Identification

Antihypertensive drugs registered in the World Health Organization Collaborating Centre (WHOCC) for Drug Statistics Methodology under the Anatomical Therapeutic Chemical (ATC) classification system were collected and their target genes were identified using the ChEMBL (*37*) (https://www.ebi.ac.uk/chembl) and DrugBank (*38*) (https://go.drugbank.com) databases (Table S4), as described previously (*10*).

#### Mendelian Randomization

An appropriate MR analysis should satisfy three key assumptions (*7*). First, the genetic variant, the so-called GI, is associated with the exposure (“relevance assumption”). Since a weak instrument bias could be a risk factor for this assumption, we calculated the variance in each gene explained by GI (R^2^) and F-statistics to assess instrumental strength (*39*). As a convention, F-statistic >10 is indicative of weak instrument bias (*40*). Second, there should be no unmeasured confounders between GI and outcome (“independence assumption”). In genetic studies, different population structures are major confounders; thus, we exclusively used data of European ancestry for exposures and outcomes to minimize genetic confounding. We also conducted multivariate MR (MVMR) and subgroup analyses to address potential confounders. Third, GI affects the outcome only through exposure to interest (“exclusion restriction”). Although vertical pleiotropy is acceptable, horizontal pleiotropy can substantially weaken the assumption of MR. We examined unbalanced pleiotropy using the Egger regression intercept and horizontal pleiotropy using a global MR-PRESSO test (*41*). We also reviewed the biological plausibility and tested potential alternative mediating pathways to exclude any pleiotropy undermining the MR results.

#### MR Analysis Using eQTL and GI Validation

We used a summary data-based MR (SMR) approach for eQTL-based MR analysis, which integrates GWAS and eQTL data (*42*). The SMR approach selects the single most significantly associated eQTL SNP (top *cis*-eQTL SNP) for each gene. Heterogeneity in dependent instruments (HEIDI) was tested using multiple SNPs in a *cis*-eQTL region to distinguish pleiotropy due to linkage disequilibrium and to determine colocalization (*42, 43*). We used the HEIDI test *P*-value threshold <0.01 to indicate an association due to a linkage scenario.

Among the 108 antihypertensive drug target genes identified in the DrugBank and ChEMBL databases, we curated 64 drug target genes with significant (*P*<5×10^−8^) eQTL SNP in at least one tissue (Table S4). We then conducted SMR using eQTL for the 64 target genes as exposure and systolic blood pressure (SBP; 757,601 individuals) (*44*) as an outcome to validate whether the instruments reflected the actual drug effects (positive control; Table S5). A total of 23 target genes whose expression was associated with SBP with at least nominal significance (*P*<0.05) were deemed valid and used for further analysis.

#### MR Analysis with Drug Target Gene Expression and AD

We performed SMR using eQTL for 23 drug target genes as exposure and AD as the outcome. To minimize false inferences, we applied a strict threshold to correct for multiple testing (Bonferroni correction threshold *P*<0.05/56 [56 overlapping genes in multiple tissues (Table S6)]). Only those passing the Bonferroni and HEIDI thresholds (*P*<0.01) were considered robust for confidentially inferring causal associations. Given that each antihypertensive drug acts as either an agonist or an antagonist of the drug targets, the odds ratio (OR) was harmonized to reflect the actual effect of the drug on target genes. Therefore, OR >1.00 indicates increased risk of disease due to antihypertensive drug use.

#### Validation for Association Between ACE and AD at Protein Level Using pQTL

Since pQTL data are relatively limited than eQTL data, we used eQTL MR for the main analysis to cover various antihypertensive drug classes, whereas we used pQTL to validate the findings from the main analysis. To observe whether different MR methods produce consistent results, we used conventional MR approaches for pQTL analysis in addition to SMR: conventional MR methods (robust MR) include inverse variance weighted, weighted median, MR-Egger, and MR-PRESSO. The significant SNPs (*P*<5×10^−8^) of protein GWAS in weak pairwise linkage disequilibrium (*r*^2^<0.1) were used as drug target gene proxies to increase the proportion of variance explained by the instrument and maximize instrument strength (*9, 10*). As done for eQTL instruments, the effect of ACE pQTL instruments on blood pressure (BP) was assessed to validate whether the instruments reflect the antihypertensive drug effect (positive control; Table S9).

#### Discovery and Validation Sets for pQTL Analysis

To strengthen this finding, we structured an analysis to discover and validate the association between ACE pQTL and AD using different datasets (Fig. S1, Table S1). For the discovery set, pQTL data of whole blood from 35,559 Icelanders, SBP data from 757,601 UK Biobank and ICBP consortium participants, and AD data from 35,274 clinically diagnosed cases (59,163 controls) were used for robust MR and SBP-adjusted MVMR. For the validation set, pQTL data of whole blood from 7,213 ARIC study participants of European ancestry and AD data from a genome-wide meta-analysis of PCG-ALZ, IGAP, ADSP, and UK Biobank (AD proxy) were used for robust MR. No noticeable sample overlap was detected between exposure and outcome or between discovery and validation sets. ACE pQTL data for the cerebrospinal fluid (CSF) were obtained for sensitivity analysis.

#### Validation for Association Between ACE and AD at Population Level Using Polygenic Risk Score

Polygenic risk score (PRS) was calculated for 338,645 unrelated, white British, and genotype quality-controlled participants with both clinical and genotype data. AD cases and diagnosis dates were ascertained using hospital inpatient records containing data from the Hospital Episode Statistics for England (HES), Scottish Morbidity Record (SMR), and Patient Episode Database for Wales (PEDW). A total of 1,756 patients with AD were identified as having primary/secondary diagnosis codes for AD from the Death Registry, International Classification of Diseases (ICD) codes 9 and 10: ICD-9 (331.0) and ICD-10 (F00.0, F00.1, F00.2, F00.9, G30.0, G30.1, G30.8, and G30.9) (*45*). The participants were followed up until the date of the first AD diagnosis, death, loss to follow-up, or the end of follow-up for the current data release (July 31, 2021, for England and Scotland; February 28, 2018, for Wales), whichever came first.

We used PRS-CS based on a high-dimensional Bayesian regression framework for the PRS calculation (*46*). Since no biobanks or large cohorts concomitantly retain both individual genotypes and laboratory data of serum ACE levels, we could not train or test for the best-fitted PRS models. Therefore, we used PRS-cs “auto” rather than selecting one PRS threshold to avoid potential overfitting issues. The omission of a training set may not be problematic because finding the best-fit PRS model is not a priority and is common practice, as shown in previous studies (*47, 48*).

The genetic variant load associated with serum ACE protein levels was computed for 338,645 individuals based on ACE GWAS of individuals of European ancestry (*35*). The participants were divided into quintiles according to their *ACE* PRS and categorized as low (lowest quintile), intermediate (quintiles 2–4), and high (highest quintile); low *ACE* PRS indicates low genetically predicted serum ACE level. The association between *ACE* PRS and AD (n=1,756) was assessed using multivariable logistic regression and Cox proportional hazards regression models. The model was adjusted for age, sex, education, smoking, alcohol consumption, depression, diabetes mellitus, hypertension, body mass index, *APOE ε4* (*APOE4*) risk allele count (0, 1, 2), genotype array, and the first 10 principal components of ancestry. A previous study reported different effects of ACE inhibitors on AD in patients carrying *APOE4* alleles (*49*); thus, we conducted a subgroup analysis of *APOE4* allele carriers versus non-carriers. Individuals with at least one *APOE4* allele were considered *APOE4* carriers. We attempted to investigate the association between ACE and *APOE4*, but no overlap of significant SNPs between the two GWAS prevented MR analysis.

#### Sensitivity Analyses

To investigate whether the significant association between a drug target gene and disease risk was mediated by BP or other pathways, we conducted pQTL MR analysis of the renin–angiotensin system (RAS) to examine the association between ACE-related pathway enzymes and AD. Moreover, MR for the association between serum ACE levels and AD- or vascular dementia (VD)-related markers (amyloid-beta [Aβ], phosphorylated tau [p-tau], total tau [t-tau], and white matter hypersensitivity [WMH]) was performed to investigate the potential pathway for the ACE effect on AD. The genotype data for AD- and VD-related markers are provided in Table S1.

To understand the relationship between BP and AD, we conducted MR analysis to determine the association between genetically predicted SBP and AD. In contrast to the lenient clumping applied for molecular exposure (1 Mb window and *r*^2^<0.1), which generally has a small number of significant SNPs, we used a conventional clumping standard for phenotypic exposure, such as SBP (10 Mb window and *r*^2^<0.01), based on the rationale provided in a previous study and referred to how they differentially clumped for a lipid-lowering agent target gene (such as HMGCR) and low-density cholesterol level (*9*).

#### Statistical Power

Since low statistical power undermines the chance of detecting a true effect and increases the likelihood of false-negative findings (*50*), we calculated the statistical power for non-significant MR results as reported previously (*51*) (http://sb452.shinyapps.io/power; Tables S18–19).

#### Statistical Analysis

In addition to the meta-analysis, all computational and statistical analyses were carried out using SMR version 1.02 and the *TwoSampleMR* package of R version 4.0.4 (R Project for Statistical Computing).

### Cohort Study

A total of 338,645 unrelated, white-British, and genotype quality-controlled participants with both clinical and genotype data were subjected to polygenic risk score (PRS) calculation. The AD cases and diagnosis dates were ascertained using hospital inpatient records containing data from the Hospital Episode Statistics for England (HES), Scottish Morbidity Record (SMR), and Patient Episode Database for Wales (PEDW). A total of 1,756 AD patients were identified as having primary/secondary diagnosis codes for AD from the Death registry, International Classification of Diseases (ICD) codes 9 and 10: ICD-9 (331.0) and ICD-10 (F00.0, F00.1, F00.2, F00.9, G30.0, G30.1, G30.8, G30.9) (*45*). Participants were followed up until the date of first AD diagnosis, death, loss to follow-up, or the end of follow-up for the current data release (July 31, 2021, for England and Scotland; February 28, 2018, for Wales), whichever came first.

We used PRS-CS based on a high-dimensional Bayesian regression framework for PRS calculation (*46*). Since there were no biobanks or large cohorts concomitantly retaining both individuals’ genotype and laboratory data of serum ACE levels, we could not train or test for best-fitted PRS models. Therefore, we used PRS-cs “auto” rather than selecting one particular PRS threshold to avoid potential overfitting issues. The omission of a training set may not be problematic because finding the best-fitted PRS model is not our priority and is a common practice, as shown in previous studies (*47, 48*).

The genetic variant load associated with serum ACE protein level was computed for 338,645 individuals based on GWAS for serum ACE protein level of individuals of European ancestry (*35*). Participants were divided into quintiles according to their ACE PRS and categorized as low (lowest quintile), intermediate (quintiles 2–4), and high (highest quintile); low ACE PRS indicates a lower genetically predicted serum ACE level. The association between the ACE PRS and AD (n = 1,756) was assessed using multivariable logistic regression and Cox proportional hazards regression models. The model was adjusted for age, sex, education, smoking, alcohol consumption, depression, diabetes mellitus, hypertension, body mass index, *APOE ε4* (*APOE4*) risk allele count (0, 1, 2), genotype array, and the first 10 principal components of ancestry. A previous study reported different effects of ACE inhibitors on AD for those carrying *APOE4* alleles (*49*), and thus, we conducted a subgroup analysis for *APOE4* allele carriers versus non-carriers. Individuals with at least one *APOE4* allele were considered *APOE4* carriers. We attempted to investigate the association between ACE and *APOE4*, but no overlapping of significant SNPs between the two GWAS prevented the MR analysis.

### Meta-Analysis of RCTs

Hughes et al. conducted the latest meta-analysis of randomized controlled trials (RCTs) investigating the effect of antihypertensive drugs on dementia (*2*), which was performed for all-cause dementia, including AD and VD, although they had distinct pathophysiologies, etiologies, and susceptibilities to atherosclerosis and cerebral hemodynamics (*52–54*). Therefore, we performed a meta-analysis of a subset of RCTs included in a previous meta-analysis (*2*) that provided isolated results from patients with AD. The analysis was performed using Review Manager statistical software (REVMAN version 5.3) under a random-effects model.

### Experimental procedures

#### Plasmid

The following plasmids were obtained from Addgene: pLVX-UbC-rtTA-Ngn2 (#127288), psPAX2 (#12260), pMD2.G (#12259), VN-Tau (#87368), and Tau-VC (#87369).

#### Animal Studies

Heterozygous Tau P301S mice (Jackson Laboratory) and wild-type littermates (C57BL6/J background) were housed under specific pathogen-free conditions with a 12-hour light/dark cycle and *ad libitum* access to food and water. At 4 months of age, P301S mice were orally administered vehicle (5% DMSO [Sigma, 41639] in PBS), ACE inhibitors (ramipril, 2.5 mg/kg; enalapril, 0.625 mg/kg), or ARB (losartan, 6.25 mg/kg) for 4 months. Behavioral assessments were performed 2 and 4 months after treatment. All experimental procedures were reviewed and approved by the Institutional Animal Care and Use Committee of Sungkyunkwan University (SKKUIACUC2023-07-35-2 and SKKUIACUC2025-04-81-1).

#### Cell Line Culture and Stable Cell Line Generation

SH-SY5Y, HEK293T, L929, and BV2 cell lines (ATCC) were cultured in Dulbecco’s Modified Eagle’s Medium (DMEM, Hyclone, SH0022.01) supplemented with 10% fetal bovine serum (FBS; Gibco, 12483020) and 1% penicillin–streptomycin (Capricone, PS-B) at 37℃ in 5% CO_2_. SH-SY5Y cells expressing VN-Tau and Tau-VC were generated by transfection, sorted by fluorescence-activated cell sorting (FACS), and maintained in G418-containing medium.

#### Lentiviral Production

Lentivirus was produced in HEK293T cells by co-transfection of 7.5 μg psPAX2, 5 μg pMD2.G, and 2 μg pLVX-UbC-rtTA-Ngn2 using PEI (Sigma, 408727). After 24 h, the medium was replaced with complete DMEM supplemented with 25 mM HEPES (Gibco, 15630080). The supernatants were harvested after 48 h, centrifuged at 800× *g* for 10 min, filtered (0.45 μm), and concentrated using Lenti-X Concentrator (Clontech, 631232).

#### Human iPSC Generation from Peripheral Blood Mononuclear Cells

Peripheral blood mononuclear cells (PBMCs) from a patient with early-onset AD and ApoE4/4 genotype (Samsung Medical Center) were maintained in QBSF-60 (Quality Biological)-based PBMC medium supplemented with 50 ng/mL SCF (Gibco, PHC2111), 10 ng/mL IL-3 (Gibco, PHC0031), 2 U/mL EPO (STEMCELL Technologies, 78007), and 40 ng/mL IGF-1 (Gibco, PHG0071). Half of the medium was replaced every alternate day. On day 4, the cells were reprogrammed using the CytoTune-iPS 2.0 Sendai Reprogramming Kit (Invitrogen, A16517) according to the manufacturer’s instructions. Three days post-transduction, the cells were transferred to Matrigel-coated dishes (Corning, 356234), and the medium was gradually transitioned to TeSR-E8 (STEMCELL Technologies, 05990). iPSCs colonies with the appropriate morphology and size were manually selected and expanded.

#### Human ESCs and Human iPSCs Culture

The *APOE3/3* iPSC line was derived from a healthy 75-year-old female individual by Dr. Yankner’s Laboratory as previously reported (*55*). Human ESCs and iPSCs were cultured in mTeSR^TM^1 medium (STEMCELL Technologies, 85850) on Matrigel-coated plates. During plating, Y-27632 (Tocris, 1254) was added to enhance cell survival. The cells were maintained at 37℃ in a humidified incubator with 5% CO_2_.

#### Generation of Ngn2-iPSCs and Neuronal Differentiation

Human iPSCs and human ESCs were transduced with lentivirus carrying rtTA and Ngn2 in the presence of 8 μg/mL polybrene (Sigma, TR-1003) and selected with 0.5 μg/mL puromycin. The differentiation protocol was adapted from that of Zhang et al. (*56*) with minor modifications. Ngn2-expressing iPSCs were plated on poly-L-ornithine (Sigma, P3655)- and laminin (Gibco, 23017015)-coated plates and cultured to 70–90% confluence (day 1). On day 2, the media were replaced with DMEM/F12 medium supplemented with 1% N-2 (Gibco, 17502048), 1% NEAA (Gibco, 11140050), and 1% penicillin/streptomycin (P/S), along with 10 ng/mL NT-3 (PeproTech, 450-03), 10 ng/mL BDNF (R&D Systems, 248-DBD), and 4 μg/mL doxycycline (Clontech, 631311). On day 4, the medium was replaced with Neurobasal medium (Gibco, 21103049) supplemented with 2% B-27 (Gibco, 17504044), 1% GlutaMAX (Gibco, 25030081), 1% P/S, 2 μM cytosine β-D-arabinofuranoside (Sigma, C1768), and the same concentrations of growth factors and doxycycline. From days 6–14, the medium was changed every other day with continuous supplementation. From day 10 onward, 2.5% FBS was added.

#### Primary Mouse Microglia Culture

Brains were isolated from C57BL/6 wild-type mice on postnatal days 1–2, as previously described (*57, 58*). Following trypsinization and mechanical dissociation, the cells were filtered through a 70-μm strainer and seeded onto T75 flasks pre-coated with poly-D-lysine in 20 mL DMEM supplemented with 10% heat-inactivated FBS and 1% P/S. After 24 h, the medium was replaced with a fresh medium. On DIV7 and DIV9, 4 mL L929 cell-conditioned medium (LCM) was added to each flask. On DIV10–11, microglia were harvested by mechanical shaking and plated (2–2.5×10⁵ cells/mL) in medium containing 10% LCM. The culture medium was replaced daily.

#### Morris Water Maze, Reversal Learning

Morris water maze test was performed as described previously (*58*). A round pool with a diameter of 120 cm was filled with water, and white non-toxic paint was added. The water temperature was maintained at 21–22°C throughout the experiment. A hidden platform was positioned 1 cm below the water surface in one of the quadrants, and distinct visual cues were placed around the pool. The test consisted of 5 consecutive days of training trials, followed by a probe trial. During the training trials, mice (6-month-old) were allowed to swim and locate the hidden platform for 60 s. If the mice failed to locate the hidden platform, they were guided to it and allowed to remain there for 10 s to observe their surroundings. The escape latency was measured for three different trials each day, with 1-h intervals between each trial. The probe trial was conducted 24 h after the last training session. During the probe trial, the hidden platform was removed from the pool, and the mice were allowed to swim freely for 60s. The number of platform location crossings were recorded and analyzed.

Mice at the age of 8 months were subjected to the second Morris water maze test, which was performed as previously described (*59*) with modifications. The test consisted of four consecutive days of training trials and one probe trial. The locations of the visual cues and hidden platform were identical to those in the first Morris water maze test. The reversal learning test was performed 24 h after the second probe trial and consisted of five training days and one probe trial. The platform was relocated to the opposite location of the original platform, while the visual cues remained unchanged. The mice were allowed to swim and locate a new hidden platform for 60 s three times a day. In the final probe trial, the new hidden platform was removed from the pool, and the mice were allowed to swim freely for 60 s. The number of new platform location crossings was recorded and analyzed.

#### Novel Object Recognition Test

The Novel object recognition (NOR) test was performed as previously described (*60*). The test was conducted in three stages: habituation, training, and testing. On the first day of the test, the mice were placed in an empty open-field box (length × width × height, 30 × 30 × 30 cm^3^) and allowed to explore and adapt to the experimental conditions. The training session was conducted the day after the habituation stage. In the training session, two similar objects were placed in the box, and the mice were allowed to explore for 10 min. Mice were subjected to a test session after resting for 3 h. In the test session, one of the objects was replaced with a novel object, and the mice were allowed to explore for 10 min. The mouse exploration patterns during the test session were recorded and analyzed.

#### Tissue Sample Preparation

Tissue samples were prepared as described previously (*61*). Briefly, mice were anesthetized with Zoletil (Virbac) and Rompun (Bayer), followed by transcranial perfusion with phosphate-buffered saline (PBS). Left hemispheres of the brain were collected and fixed in 4% Paraformaldehyde (PFA) at 4°C overnight. Brain tissues were sequentially infiltrated with 10%, 20%, and 30% sucrose solutions in PBS, each step carried out at 4°C with overnight incubation. Fully infiltrated brain tissue was cryoembedded in a Tissue-Tek Cryomold (Sakura Finetek, USA) for immunohistochemical analysis. The hippocampus and cortex isolated from the right hemisphere of the brain were flash-frozen in liquid nitrogen and stored at −80°C for biochemical analyses.

#### Immunoblotting

Protein samples were prepared as described previously (*62*). Brain tissues were homogenized on ice in cold PBS using a Biomasher (Funakoshi). Brain tissues and cells used for western blotting were lysed in 10× RIPA lysis buffer (Merck Millipore, 20-188) supplemented with a phosphatase/protease inhibitor cocktail (Thermo Fisher Scientific, 78444). Lysates were incubated on ice for 20 min followed by centrifugation at 13,000 rpm for 20 min at 4°C. The resulting supernatant was used for further analysis. Protein concentrations were quantified using the Pierce BCA Assay (Thermo Fisher Scientific, 23225). Equal amounts of protein were mixed with NuPAGE LDS 4X Sample Buffer (Thermo Fisher Scientific, NP0008) containing 5% 2-Mercaptoethanol (Sigma-Aldrich, M6250) and heated for 10 min at 95°C. For the detection of high-molecular-weight tau proteins, the samples were mixed with a sample buffer without 2-mercaptoethanol. Samples were separated by SDS-PAGE (Bio-Rad Laboratories) and transferred to 0.45 μm polyvinylidene difluoride (PVDF) membranes (Merck Millipore). The membranes were blocked with 5% bovine serum albumin (Bovogen, BSA S 0.1) for 1 h at room temperature and incubated overnight at 4°C with primary antibodies. Antibodies used were as follows: anti-Tau (Tau-13; Biolegend, 834201,1:10000), anti-pTau(Ser202, Thr205) (AT8; Thermo Fisher Scientific, MN1020, 1:500), anti-pTau(Thr212, Ser214) (AT100; Thermo Fisher Scientific, MN1060, 1:500), anti-pTau(Thr181) (AT270; Thermo Fisher Scientific, MN1050, 1:500), anti-pTau(Ser262, Thr263) (Abcam, ab92627,1:500), anti-GAPDH (Novus, NB300-221, 1:1000), and anti-β-Actin (Sigma-Aldrich, A5441, 1:20000). The membranes were washed thrice with Tris-buffered saline (TBS) containing 0.1% Tween 20 (TBS-T) and incubated with peroxidase-conjugated anti-rabbit or anti-mouse secondary antibodies (Invitrogen) for 1 h at room temperature. Immunoreactive bands were detected using the Dyne ECL STAR Western blot Detection Kit (Thermo Fisher Scientific, 32106).

#### Quantitative Real-time PCR

Total RNA extraction and Quantitative real-time PCR (qRT-PCR) were performed as described previously (*61*). Briefly, total RNA was extracted from mouse cortices and cultured cells using RNAiso Plus (TaKaRa, 9108) according to the manufacturer’s instructions. The extracted RNA was reverse-transcribed into complementary DNA (cDNA) using the PrimeScript RT Master Mix (TaKaRa, RR047A). RT-qPCR was performed using the TB Green Premix Ex Taq II (TaKaRa, RR820A). All primers used for qPCR in this study are listed in Table S21.

#### Immunofluorescence

Immunohistochemical analysis was performed as previously described (*61*). Cryo-embedded brain tissues were cut into 30 μm sections using a cryostat (Leica), blocked with 2.5% donkey serum in 0.3% Triton X-100 in DPBS (PBS-T), and overnight incubated at 4°C with the following primary antibodies: anti-pTau(Ser202, Thr205) (AT8; Thermo Fisher Scientific, 1:500), anti-pTau(Thr212, Ser214) (AT100; Thermo Fisher Scientific, 1:500), anti-Iba1 (Wako, 019-19741, 1:800), and anti-GFAP (Dako, Z0334, 1:1000). The sections were then washed three times with PBS-T and incubated with a secondary antibody for 1 h at room temperature. The brain sections were mounted using an antifade mounting medium containing DAPI (Cat. No. H-1200-10). Confocal imaging was performed using Leica STELLARIS 5 (Leica).

For immunocytochemical analysis, the cells were seeded on Matrigel (Corning, NY, USA)-coated coverslips. The next day, the cells were washed three times with Dulbecco’s Phosphate-Buffered Saline (DPBS; Sigma, D5652) and fixed with 4% PFA for 10 min at room temperature. The cells were then permeabilized with 0.1% PBS-T for 5 min at room temperature and washed with DPBS at each step. The cells were blocked with 3% BSA in PBS-T for 1 h at room temperature, followed by overnight incubation at 4°C with the following primary antibodies: SSEA4 (Merck Millipore, MAB4304, 1:500), OCT4 (Cell Signaling Technology, 2750, 1:500), TRA-1-60 (Merck Millipore, MAB4360, 1:500), and Nanog (Abcam, ab80892, 1:500). The following day, the cells were washed thrice with PBS-T and incubated with secondary antibodies for 1 h at room temperature. The coverslips were mounted using an antifade mounting medium containing DAPI. Images were acquired using a Leica STELLARIS 5 and Cytation5 (BioTek).

#### Imaging Analysis

Confocal z-stack images (10 μm depth, 1 μm interval) were acquired and analyzed using ImageJ and Imaris software (Bitplane, v10.0.0). Iba1- and GFAP-positive areas were measured using ImageJ software. Volumetric analyses of Iba1-positive microglia and GFAP-positive astrocytes were performed using 3D surface rendering in Imaris.

#### RNA Sequencing

Total RNA was sent to Novogene Co., LTD (Beijing, China) for library preparation (NEBNext Ultra RNA Library Prep Kit, Illumina) and sequencing (NovaSeq 6000). Reads were filtered, trimmed, and aligned to the mouse reference genome (GRCm39, Gencode vM32) using STAR (v2.7.10a) (*63*), and gene-level counts were quantified using RSEM (v1.2.25) (*64*). Differential expression analysis was conducted using DESeq2 (v1.42.1) (*65*) with thresholds of *P*<0.05 and fold change >1.3. Heatmaps and volcano plots were generated using ComplexHeatmap (v2.18.0) and ggplot2 (v3.5.0), respectively. Gene enrichment analysis was performed using the DAVID, for Annotation, Visualization and Integrated Discovery (https://david.ncifcrf.gov).

#### Statistical Analysis

Data are presented as mean ± standard deviation (SD) or mean ± standard error of the mean (SEM), as indicated in figure legends. Statistical significance was determined using two-tailed Student’s t-tests or one-way ANOVA followed by Tukey’s or Dunnett’s post-hoc tests, as appropriate. All analyses were performed using the GraphPad Prism 10 software (GraphPad Software). *P*<0.05 were considered statistically significant (\**P*<0.05, \*\**P*<0.01, \*\*\**P*<0.001, \*\*\*\**P*<0.0001).

## Data Availability

All data produced in the present study are available upon reasonable request to the authors

## Acknowledgments

We acknowledge the participants and investigators of the UK Biobank. We sincerely appreciate Dr. Yankner at Harvard Medical School for generously providing the ApoE3 iPSC line.

## Contributors

Concept: Min Seo Kim, Dong-Gyu Jo, Hong-Hee Won

Design: Min Seo Kim, Dong-Gyu Jo, Hong-Hee Won

Data Acquisition: Woong-Yang Park, Hong-Hee Won, Dong-Gyu Jo, Hee Jin Kim, Hyo Jin Son, Kwangsik Nho, Ju-Young Shin

Computational and statistical analysis: Min Seo Kim, Minku Song, Seonghoon Hwang, Ju-Young Shin Experiments: Sunyoung Park, Jong Ho Kim, Eunae Kim

Interpretation of data: Min Seo Kim, Sunyoung Park, Jong Ho Kim, Woojae Myung, Kwangsik Nho, Zhi Yu, Pradeep Natarajan, Hee Jin Kim, Woong-Yang Park, Carlos Cruchaga, Patrick T. Ellinor, Dong-Gyu Jo, and Hong-Hee Won

Drafting of the manuscript: Min Seo Kim, Sunyoung Park, Dong-Gyu Jo, Hong-Hee Won

Critical revision of the manuscript for important intellectual content: Woojae Myung, Kwangsik Nho, Zhi Yu, Pradeep Natarajan, Hee Jin Kim, Carlos Cruchaga, Woong-Yang Park, Akl C Fahed, Yang Sui, Muhammad Ali, Katherine Gong, Patrick T. Ellinor, Dong Keon Yon, and Hong-Hee Won

Statistical supervision: Hong-Hee Won, Minku Song, Ron Do, Pradeep Natarajan, Zhi Yu

Obtained funding: Hong-Hee Won, Dong-Gyu Jo

Administrative, technical, or material support: Woong-Yang Park, Kwangsik Nho, Jinsoo Seo, and Hong-Hee Won

Supervision: Hong-Hee Won, Dong-Gyu Jo, Min Seo Kim

This manuscript has been reviewed and approved by all authors.

## Data and materials availability

All data are available in the main text or the supplementary materials. Additional data related to this paper may be requested from the authors.

## Supplementary Materials

Materials and Methods

Supplementary Text

Figs. S1 to S11

Tables S1 to S20

## FIGURE LEGENDS

**Supplementary Fig. 1.**
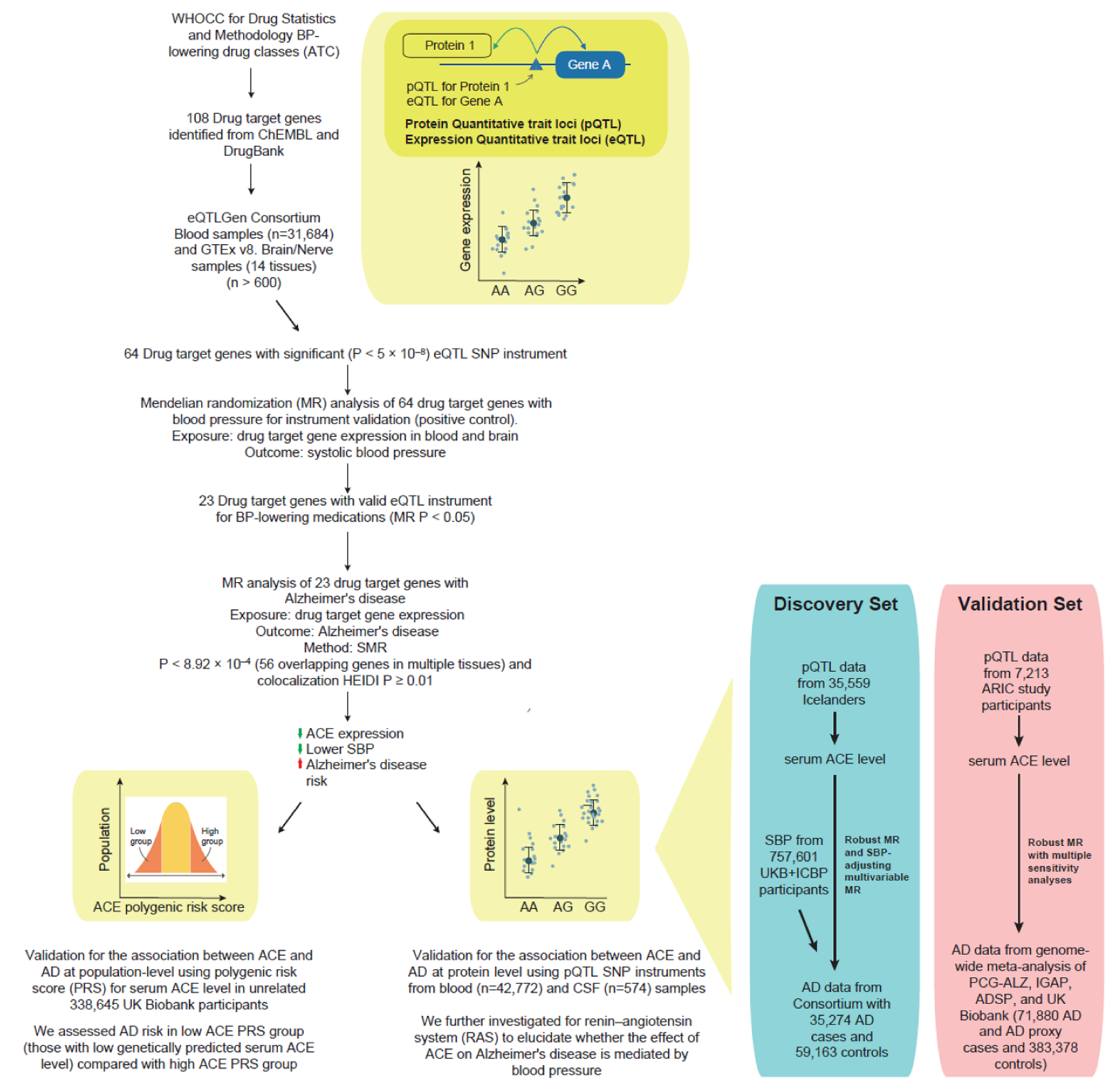
Overview of study design and key findings from the human genetics analysis. WHOCC, World Health Organization Collaborating Centre; BP, blood pressure; ATC, anatomical therapeutic chemical classification system; eQTL, expression quantitative trait loci; pQTL, protein quantitative trait loci; SNP, single-nucleotide polymorphism; SMR, summary data-based Mendelian randomization; ACE, angiotensin-converting enzyme; SBP, systolic blood pressure; CSF, cerebrospinal fluid; AD, Alzheimer’s disease.

**Supplementary Fig. 2.**
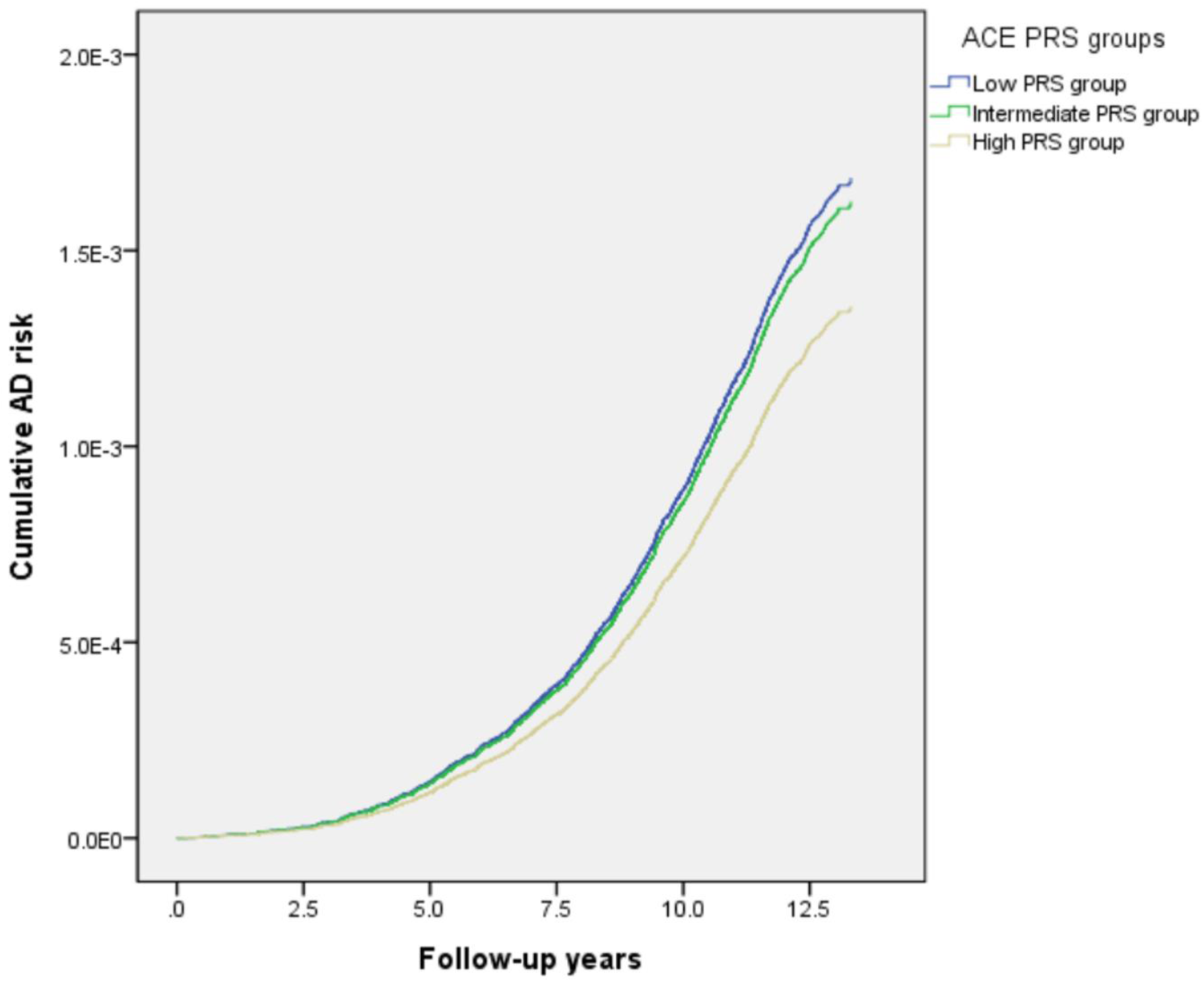
Cumulative Alzheimer’s disease risk per ACE PRS groups in UK Biobank participants during a 15-year follow-up period.

**Supplementary Fig. 3.**
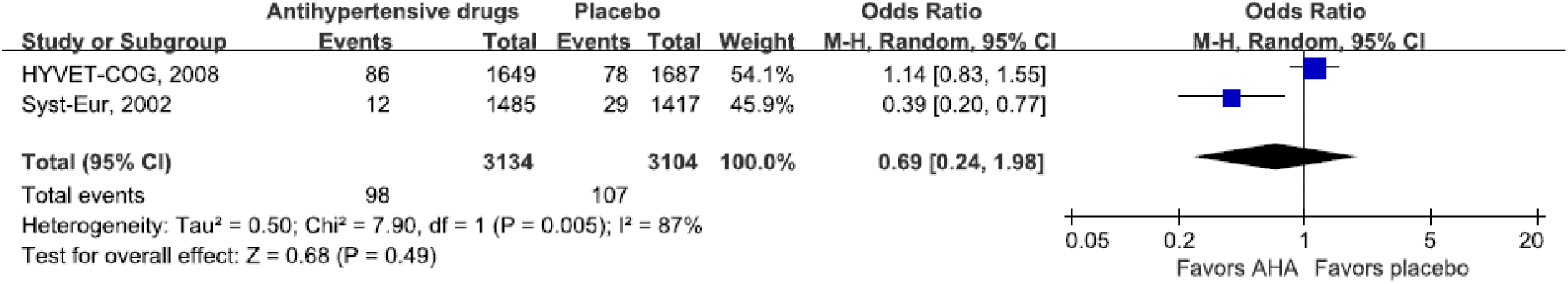
Reanalysis of the meta-analysis by Hughes et al. by incorporating only Alzheimer’s disease studies. Hughes et al. conducted meta-analysis for the association between antihypertensive agents and all-cause dementia (including AD, VD, etc.). AHA: Antihypertensive agents.

**Supplementary Fig. 4.**
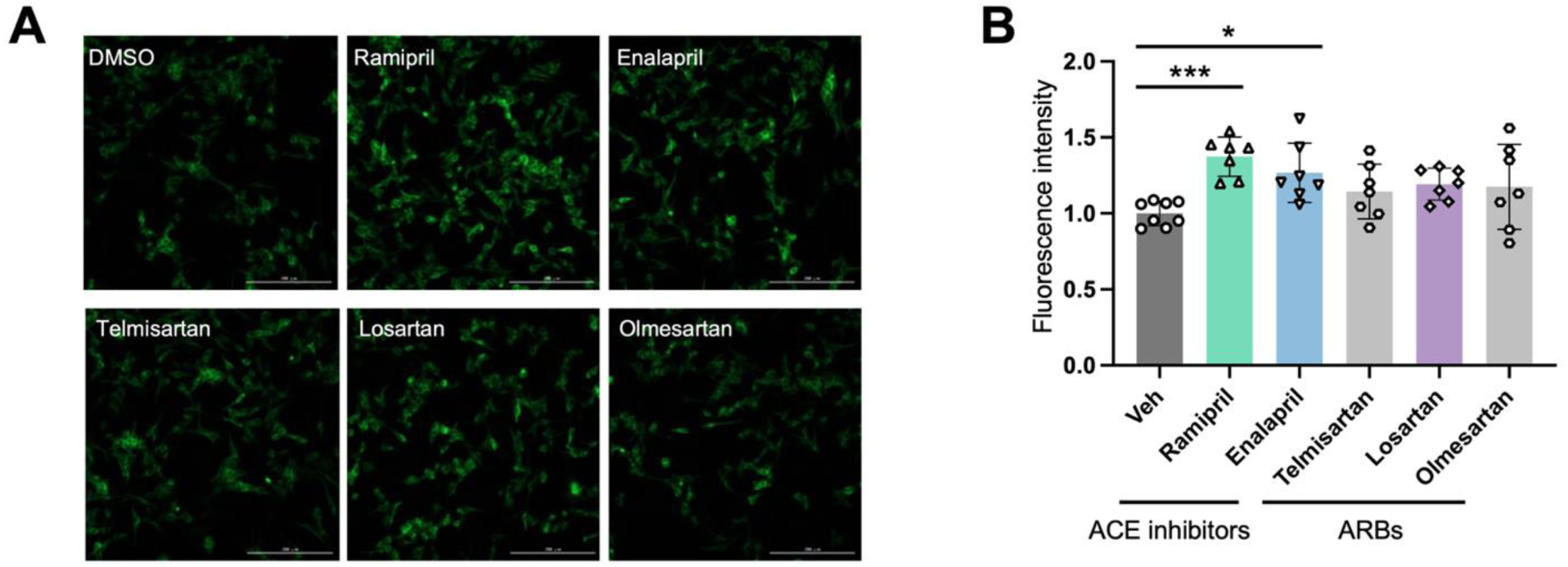
Analysis of tau aggregation in Tau-BiFC cells following treatment with ACE inhibitors and ARBs. (**A**) Representative images of Tau-BiFC cells treated with ACE inhibitors or ARBs. (**B**) Quantification of tau aggregation based on YFP signal intensity. ACE inhibitors: ramipril, and enalapril. ARBs: telmisartan, losartan, and olmesartan. Data represent mean ± SD. Statistical significance was determined using one-way ANOVA followed by Dunnett’s multiple comparisons test against vehicle-treated controls (*P < 0.05, ***P < 0.001). Scale bar, 100µm.

**Supplementary Fig. 5.**
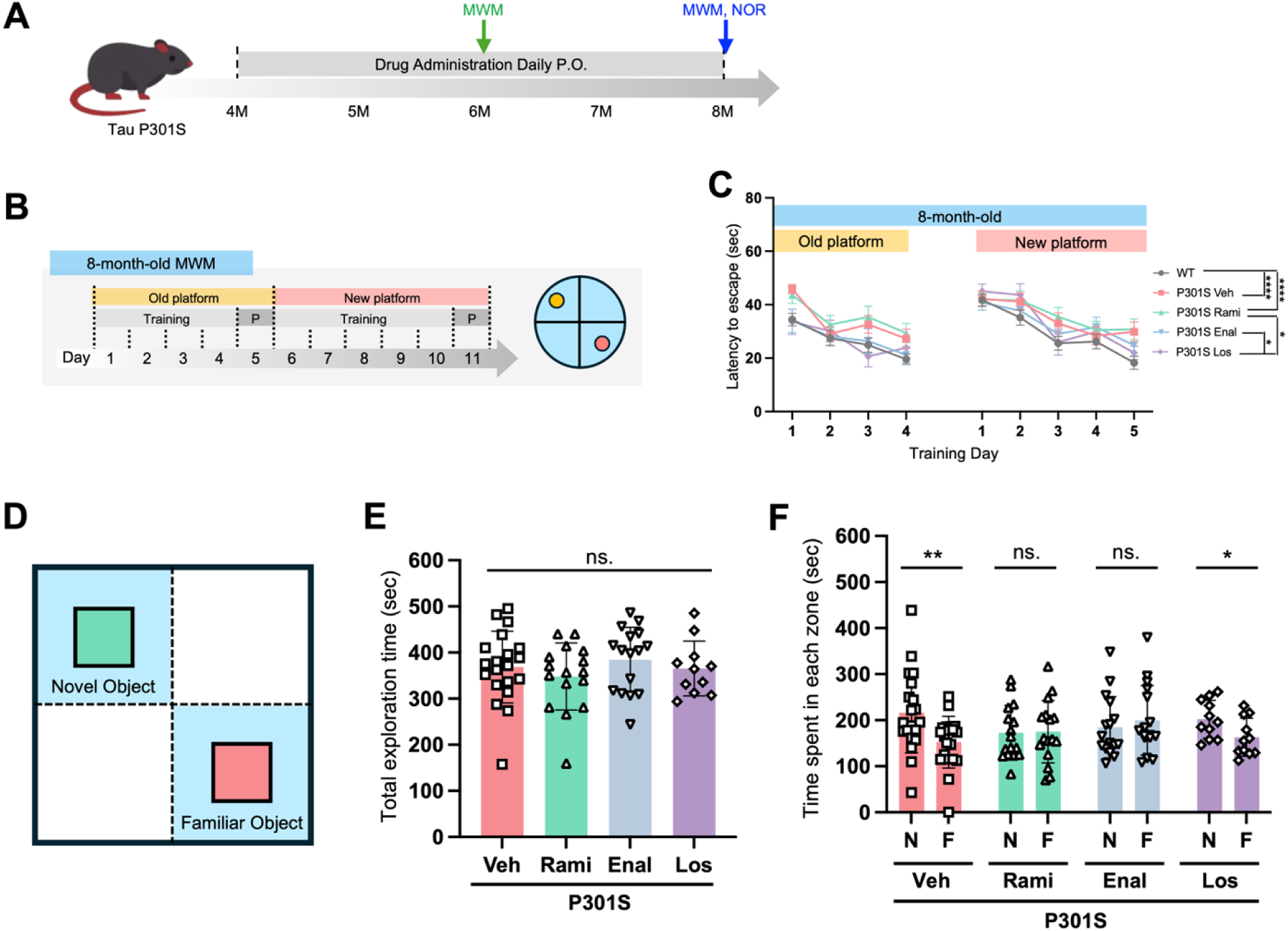
ACE inhibitors impair cognitive memory in P301S mice. **A, B**) Schematic representation of experimental design. ACE inhibitors (Ramipril, Enalapril) and an angiotensin receptor blocker (ARB; Losartan) were administered via oral injection. Group sizes: WT (n=30), P301S Vehicle (n=18), P301S Ramipril (Rami; n=16), P301S Enalapril (Enal; n=13), P301S Losartan (Los; n=10). (**C**) Escape latency during training days in the Morris water maze (MWM) test at 8 months of age. (**D**) Schematic representation of the novel object recognition (NOR) test design. Group sizes were as follows: P301S + vehicle (n = 20), P301S + ramipril (Rami; n = 16), P301S + enalapril (Enal; n = 16), P301S + losartan (Los; n = 11). (**E**) Total exploration time (seconds) spent across both object zones during the test phase. (**F**) Comparison of exploration time spent in zones containing either familiar (F) or novel (N) objects. (C) Statistical analyses were conducted using two-way ANOVA with mean ± SEM (*P < 0.05, ****P < 0.0001); (E-F) Data represent mean ± SD. Statistical analyses were conducted using one-way ANOVA followed by Dunnett’s multiple comparisons test against vehicle-treated controls in (**E**), and paired two-tailed Student’s t-test between object zones within each group in (**F**) (*P < 0.05, **P < 0.01). Veh, Vehicle; Rami, Ramipril; Enal, Enalapril; Los, Losartan.

**Supplementary Fig. 6.**
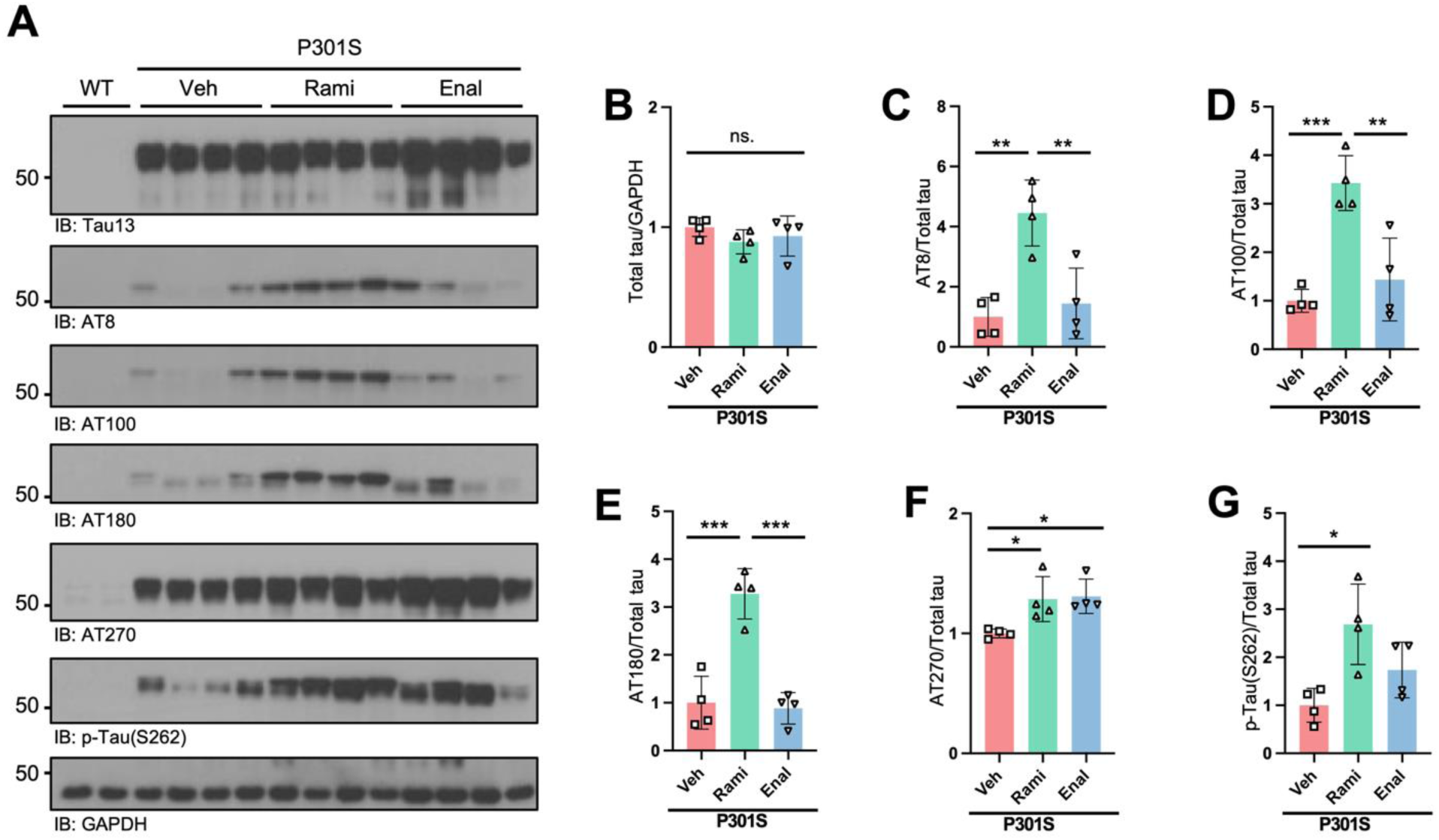
ACE inhibition increases tau phosphorylation in cortical tissues of P301S mice. (**A**) Representative immunoblots showing levels of total tau and phosphorylated tau (p-Tau) in cortical lysates. (F–K) Quantification of immunoblots: (**B**) Total tau (Tau13), (**C**) p-Tau (AT8), (**D**) p-Tau (AT180), (**E**) p-Tau (AT100), (**F**) p-Tau (AT270), (**G**) p-Tau (S262). Data represent mean ± SD. Statistical significance was assessed by one-way ANOVA followed by Tukey’s multiple comparisons test (*P < 0.05, **P < 0.01, ***P < 0.001, ****P < 0.0001). Veh, Vehicle; Rami, Ramipril; Enal, Enalapril; Los, Losartan.

**Supplementary Fig. 7.**
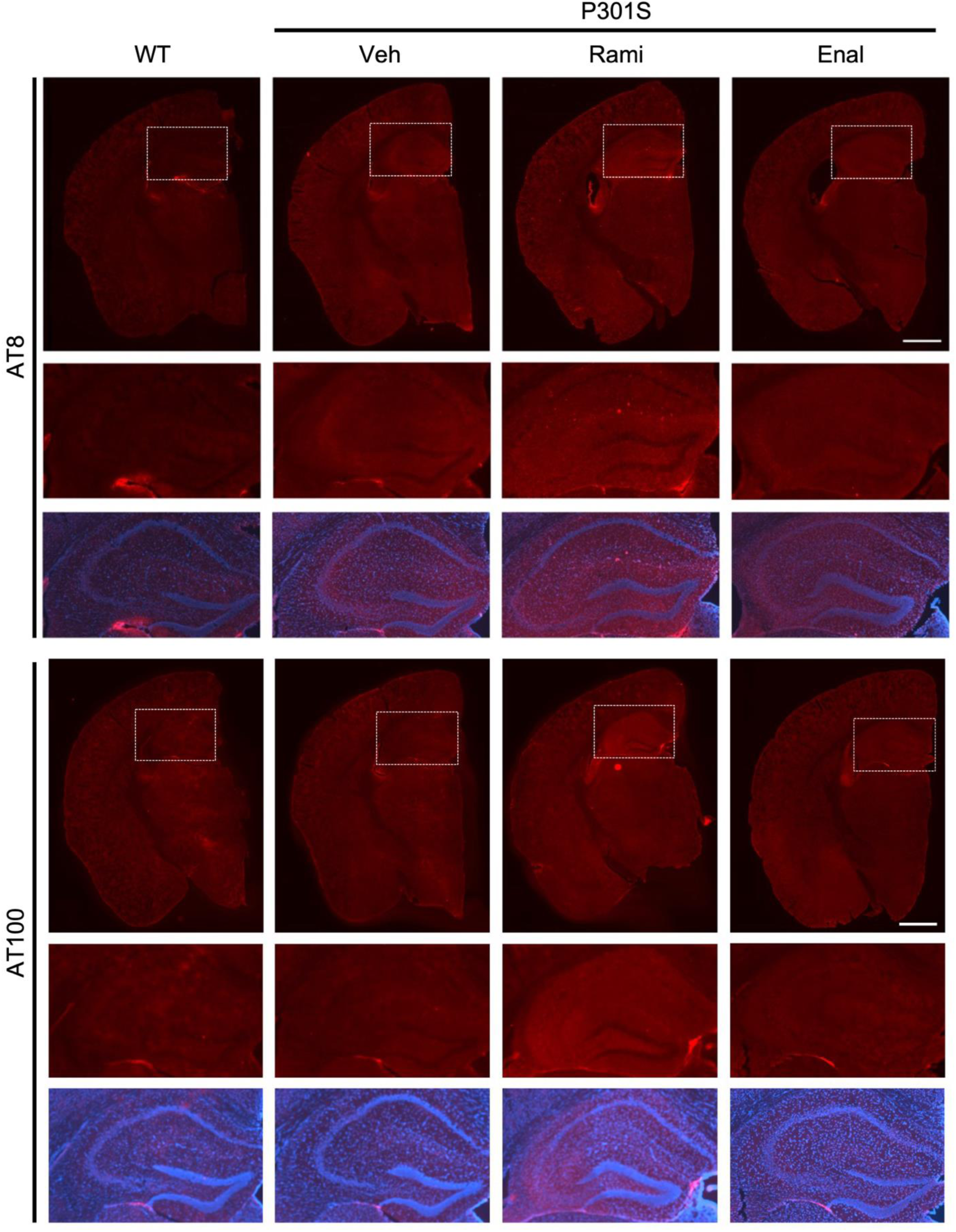
ACE inhibitors exacerbate tau pathology in P301S mice. Representative immunohistochemical (IHC) images of brain hemispheres and hippocampal regions from WT, P301S Vehicle, P301S Ramipril (Rami), and P301S Enalapril (Enal) mice. Brain sections were stained with antibodies against phosphorylated tau (AT8, upper panels; AT100, lower panels). Hemisphere-level images (top row) and magnified hippocampal views (middle row; corresponding to boxed area) are shown. Bottom-row images display merged staining with DAPI, highlighting nuclear localization and hippocampal architecture. Scale bar, 500 μm.

**Supplementary Fig. 8.**
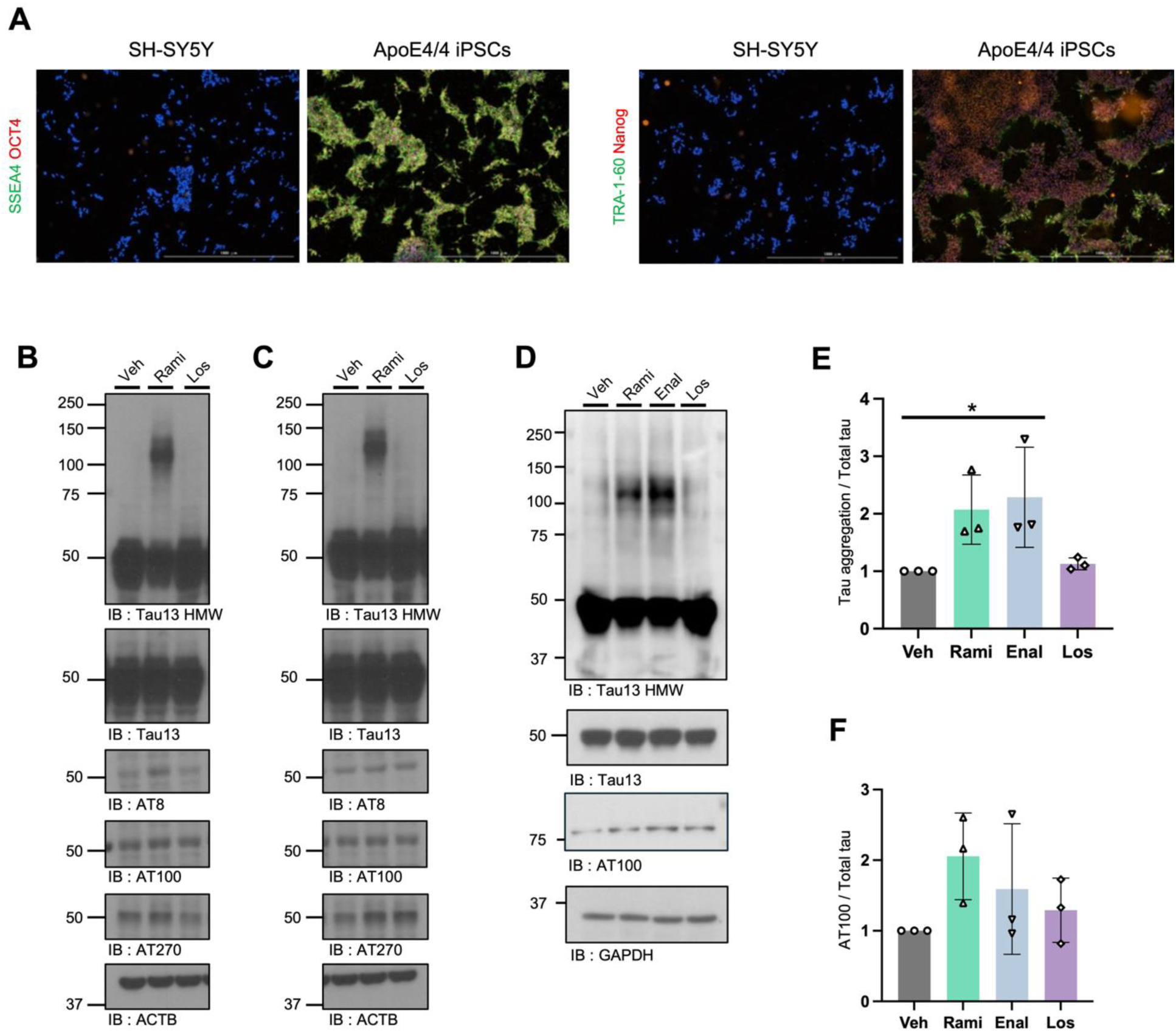
Characterization of iPSC pluripotency and tau pathology in iPSC-derived cortical neurons. (**A**) Immunofluorescence analysis of human iPSCs and SH-SY5Y neuroblastoma cells, showing specific expression of pluripotency markers (SSEA4, OCT4, TRA-1-60, and Nanog) in iPSCs. Scale bar, 1000µm. (**B** and **C**) Immunoblot analysis of total tau and phosphorylated tau in iPSC-derived cortical neurons using AT8, AT100, and AT270 antibodies. (**B**) Neurons derived from APOE3/3 iPSCs; (**C**) neurons derived from APOE4/4 iPSCs. (**D**) Immunoblot analysis of total and phosphorylated tau in neurons derived from human ESCs. (**E** and **F**) Quantification of immunoblots: (**E**) high-molecular-weight tau, (**G**) AT100. Data are presented as mean ± SD. Statistical significance was determined by one-way ANOVA followed by Dunnett’s multiple comparisons test against vehicle-treated controls (*P < 0.05). Veh, Vehicle; Rami, Ramipril; Enal, Enalapril; Los, Losartan. Scale bar, 100µm.

**Supplementary Fig. 9.**
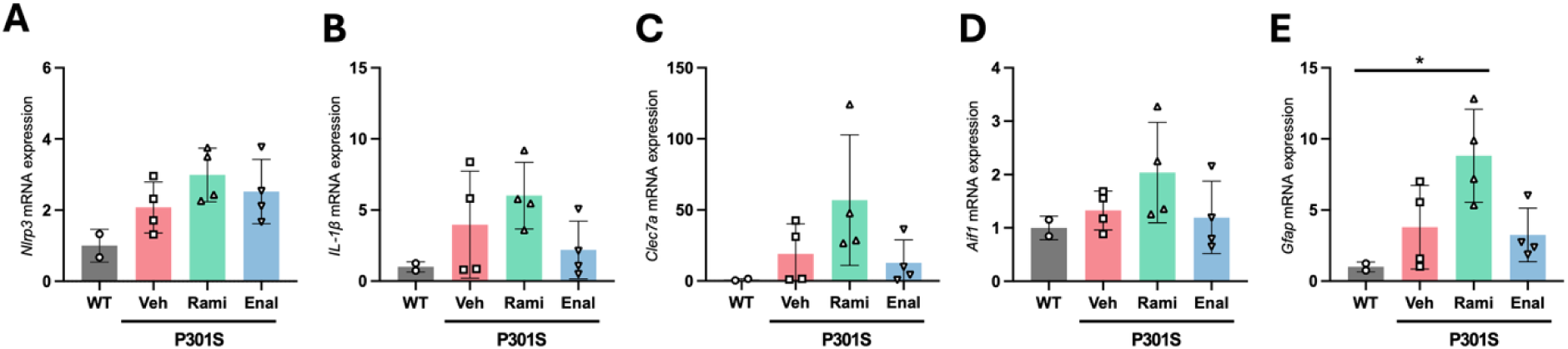
ACE inhibitors modulate expression of inflammatory genes in cortical tissue from P301S mice. (**A–E**) mRNA expression levels of inflammatory markers in cortical lysates from mice treated with ACE inhibitors: (**A**) *Nlrp3*, (**B**) *Il1b*, (**C**) *Clec7a*, (**D**) *Aif1* (Iba1), (**E**) *Gfap*. Data are presented as mean ± SD. Statistical significance was determined using one-way ANOVA followed by Tukey’s multiple comparisons test (*P < 0.05). Rami, Ramipril; Enal, Enalapril; Los, Losartan.

**Supplementary Fig. 10.**
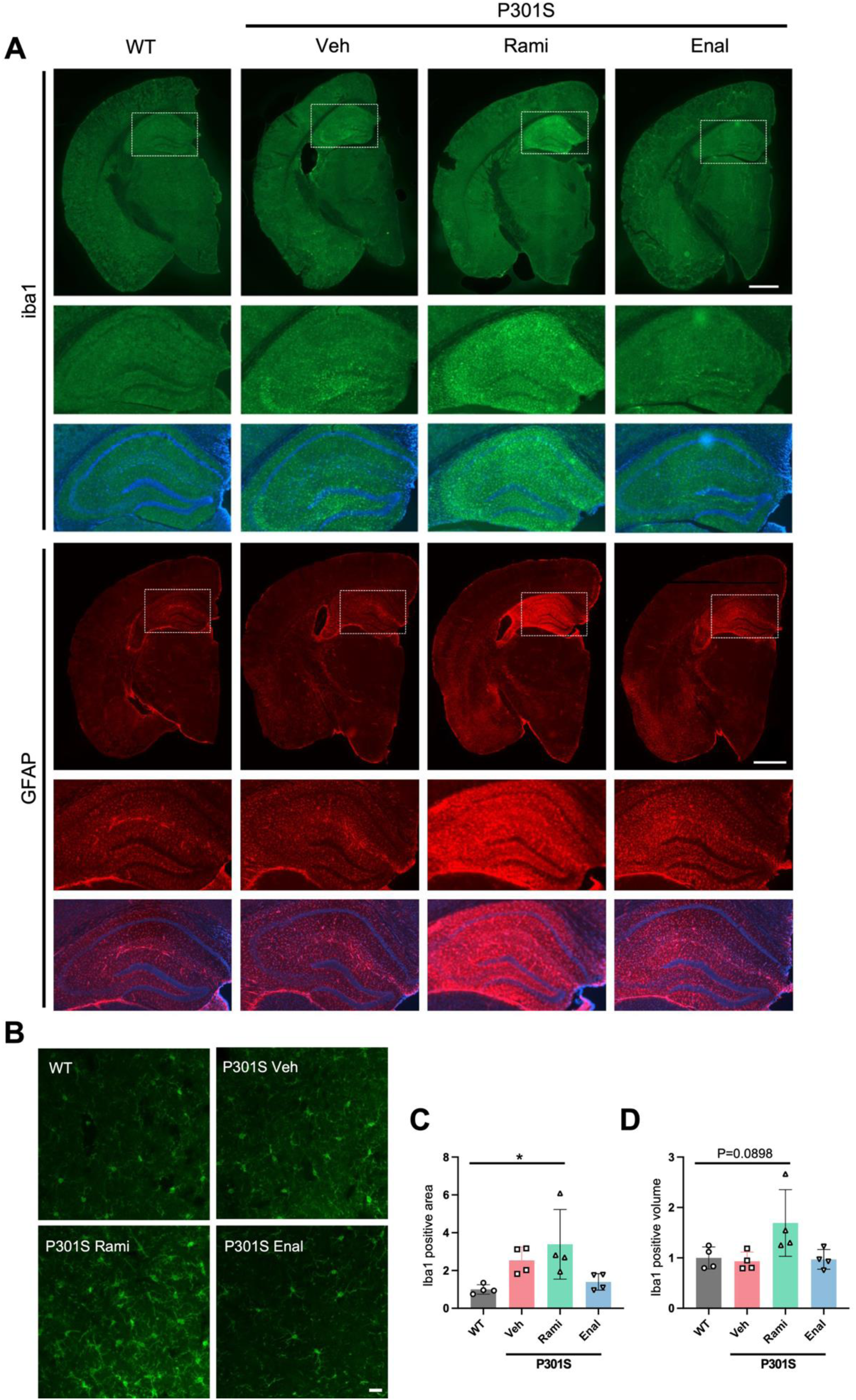
ACE inhibitors induce glial activation in P301S mice. (**A**) Representative IHC images of brain hemispheres and hippocampal regions from WT, P301S Vehicle (Veh), P301S Ramipril (Rami), and P301S Enalapril (Enal) mice. Sections were stained with Iba1 (upper panels) or GFAP (lower panels) antibodies. Hemisphere-level images (top row) and magnified hippocampal views (middle row; corresponding to boxed area) are shown for each condition with DAPI-merged images (bottom low) to visualize nuclear localization and hippocampal cytoarchitecture. Scale bar, 500 μm. (**B**) IHC images of cortical sections stained with Iba1 to assess microglial activation. Scale bar, 5 μm. (**C** and **D**) Quantification of (**C**) Iba1-positive area and (**D**) Iba1-positive volume. Data are presented as mean ± SD. Statistical significance was determined by one-way ANOVA followed by Tukey’s multiple comparisons test (*P < 0.05).

**Supplementary Fig. 11.**
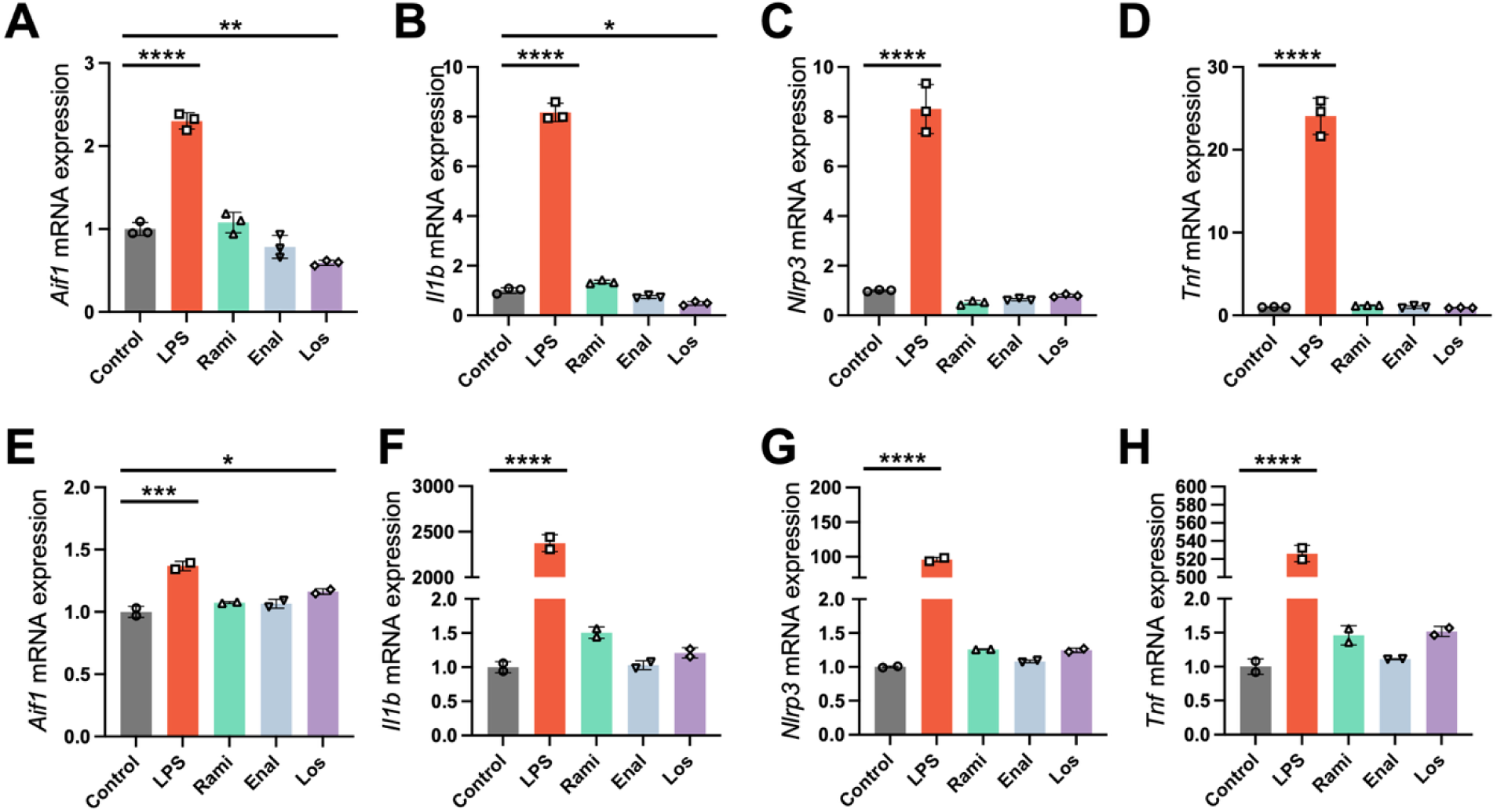
ACE inhibitors and ARBs modulate inflammatory gene expression in BV2 and primary mouse microglial cells. (**A–E**) mRNA expression levels of inflammatory markers in BV2 microglial cells treated with ACE inhibitors or ARBs: (**A**) *Aif1* (Iba1), (**B**) *Il1b*, (**C**) *Il6*, (**D**) *Nlrp3*, and (**E**) *Tnf*. (**F–J**) mRNA expression levels of inflammatory markers in primary mouse microglial cells treated with ACE inhibitors or ARBs: (**F**) *Aif1* (Iba1), (**G**) *Il1b*, (**H**) *Il6*, (**I**) *Nlrp3*, and (**J**) *Tnf*. Data are presented as mean ± SD. Statistical significance was determined using one-way ANOVA followed by Dunnett’s multiple comparisons test (*P < 0.05, **P < 0.01, ****P < 0.0001). Rami, Ramipril; Enal, Enalapril; Los, Losartan.

